# Immunogenicity, reactogenicity, and IgE-mediated immune responses of a mixed whole-cell and acellular pertussis vaccine schedule in Australian infants: a randomised, double-blind, non-inferiority trial

**DOI:** 10.1101/2023.12.20.23300336

**Authors:** Gladymar Pérez Chacón, Marie J Estcourt, James Totterdell, Julie A Marsh, Kirsten P Perrett, Dianne E Campbell, Nicholas Wood, Michael Gold, Claire S Waddington, Michael O’ Sullivan, Sonia McAlister, Nigel Curtis, Mark Jones, Peter B McIntyre, Patrick G Holt, Peter C Richmond, Tom Snelling

**Author notes:** Corresponding author: Prof Tom Snelling – Head, Health and Clinical Analytics Lab, Sydney School of Public Health, Faculty of Medicine and Health, The University of Sydney, Australia. Address: Rm 123, Edward Ford Building A27, The University of Sydney NSW (Australia), 2006.

## Abstract

**Background:** In most high-income countries, infant vaccination with acellular pertussis (aP) vaccines is the standard of care for the prevention of pertussis disease. Based on immunological and epidemiological evidence, we hypothesised that substitution of the first aP dose in the vaccination schedule with whole-cell (wP) vaccine might protect against the development of IgE-mediated food allergy. Here we report the results of a randomised comparison of the reactogenicity, immunogenicity, and IgE-mediated immune responses of a mixed wP/aP primary schedule versus the standard aP-only schedule.

**Methods:** OPTIMUM is a Bayesian, two-stage, double-blind, group sequential trial, enrolling healthy Australian-born infants. At approximately 6 weeks old, participants are randomly assigned (1:1) to a first dose of a pentavalent wP combination vaccine (DTwP-Hib-HepB; Pentabio, PT Bio Farma, Indonesia) versus a hexavalent aP vaccine, which includes inactivated poliovirus vaccine (IPV) types 1, 2, and 3 in its formulation (DTaP-Hib-HepB-IPV; Infanrix Hexa, GlaxoSmithKline, Australia). All infants receive a hexavalent aP vaccine at 4 and 6 months old, as well as a tetravalent aP-based formulation at 18 months old (DTaP-IPV; Infanrix-IPV, GlaxoSmithKline, Australia) to ensure at least three doses of IPV as part of their childhood schedule while preserving blinding. In stage one, pertussis antigen-specific IgG responses were measured before and approximately one month after the 6-month aP vaccine doses. The immunogenicity of the mixed schedule (wP/aP/aP) was defined as being non-inferior to that of the aP-only schedule (aP/aP/aP) using a non-inferiority margin of 2/3 on the ratio of the geometric mean concentrations of pertussis toxin (PT) IgG approximately one month after the 6-month aP dose. Solicited adverse reactions were summarised by study arm and included all children who received the first dose of either wP or aP. Parental acceptance was assessed using a 5-point Likert scale. The trial is registered with ANZCTR (ACTRN12617000065392p).

**Results:** Between March 7, 2018 and January 13, 2020, 150 infants were randomised (75 per arm). Demographic and baseline characteristics were balanced across the study arms. Most infants were born to aP-vaccinated mothers. In the intent-to-treat analysis, PT-IgG responses of the mixed schedule were non-inferior to the aP-only schedule approximately one month after a 6-month aP dose [geometric mean ratio (GMR) = 0·98, 95% Bayesian credible interval (0·77 to 1·26); probability (GMR > 2/3) > 0·99]. Severe solicited systemic adverse reactions were reported among 14 of 74 (19%) infants after a first dose of wP and among 8 of 72 (11%) infants after a first dose of aP; irritability after the first dose of pertussis-containing vaccine was the most frequent severe event (11 of 74 [15%] wP recipients versus 7 of 72 [10%] aP recipients). Within 6 months of enrolment, 7 serious adverse events were reported, with none deemed related to the study vaccines. Parental acceptance of the mixed schedule was high (97% would agree to have this schedule again).

**Interpretation:** The mixed wP/aP schedule was associated with more reactions than the aP-only schedule, but these were mostly non-severe. The mixed schedule was well accepted by parents and evoked non-inferior PT-IgG responses after completion of the three-dose primary series.

**Funding:** Telethon New Children’s Hospital Research Fund and National Health and Medicine Research Council.

**Research in context:** We searched PubMed on April 17, 2023, for paediatric studies of heterologous priming with whole-cell pertussis (wP) vaccine and acellular pertussis (aP) vaccine, with no language or date restrictions. We used the terms [“whole-cell pertussis vaccine” AND “acellular pertussis vaccine”], OR [“IgE” AND “tetanus toxoid”]. Of the 997 articles retrieved, we found no published randomised comparisons between heterologous versus exclusive primary routine vaccination with either wP or aP-based formulations.

In two observational studies, laboratory-confirmed pertussis disease was less common among school-aged children and adolescents who received wP versus aP as a first dose. A heterologous wP/aP primary schedule (in which the first dose was wP) was associated with lower rates of pertussis disease than an aP-only primary schedule.

In a case-control study, pertussis was less common among children who had received mixed wP/three-component (3c)-aP vaccine (including pertussis toxoid, filamentous haemagglutinin, and pertactin) versus those exclusively primed with 3c-aP vaccine formulations. No evidence of a difference was observed among those vaccinated with a heterologous wP/five-component (5c)-aP primary schedule versus those exclusively primed with a 5c-aP vaccine formulation (including the above-mentioned pertussis antigens as well as fimbriae type 2 and 3). In contrast, another case-control study found that compared to the 5c-aP-only priming strategy, a primary series including one or more doses of a wP vaccine formulation, with reported efficacy against laboratory-confirmed pertussis between 36% (95% CI 14% to 52%) and 48% (95% 37% to 58%), was associated with higher vaccine effectiveness against pertussis disease more than a decade after priming. In none of the case-control analyses was the nature of the heterologous schedules further described.

Two clinical and immunological studies reported that wP-only schedules were associated with lower post-priming tetanus toxoid (TT)-IgE concentrations than homologous priming with aP-containing vaccines. An additional study reported lower TT-IgE concentrations after a first dose of wP versus aP.

**Added value of this study:** To our knowledge, this is the first trial to evaluate the immunogenicity, reactogenicity, and IgE-mediated immune responses to a mixed primary schedule consisting of a first dose of wP given at approximately 6 weeks old, followed by aP at 4, and 6 months old.

**Implications of all the available evidence:** This trial provides supporting evidence of the safety and immunogenicity of a mixed wP/aP vaccine schedule in a setting with high maternal pertussis vaccine coverage. The findings warrant further investigation of the comparative clinical effects of a mixed wP/aP versus the standard aP-only schedule.

## Introduction

Whole-cell pertussis (wP) vaccines cause adverse reactions in infants frequently, but these are generally benign and self-limited.^1^ Since the early 1990s, wP vaccine formulations have been discontinued from use in most high- and some middle-income countries and replaced by less reactogenic subunit acellular (aP) vaccines.^2^ Data from pertussis outbreaks suggest that primary schedules comprising at least one wP vaccine dose provide better long-term protection than aP-only primary schedules.^3,4^

We have previously reported that compared to matched population controls, Australian children with IgE-mediated food allergy were less likely to have received wP rather than aP as their first pertussis vaccine dose in infancy (odds ratio [OR] 0·77; 95% CI 0·62 to 0·95).^5^ Consistent with this observation, previous studies have described differences in the immune effects arising from vaccination with wP versus aP vaccines in infancy. Compared to exclusive vaccination with aP which elicits a Th_2_-polarised immunophenotype, a first dose of wP induces relative Th_1_/Th_17_ immune polarisation, reducing the production of aP antigen-specific type 2 cytokines and down-regulating the synthesis of total IgE, and specific IgE responses against tetanus toxoid (TT), diphtheria toxoid (DT), pertussis toxin (PT), and food antigens.^6–11^ We therefore hypothesise that mixed priming using a first dose of wP followed by aP vaccine doses could protect against IgE-mediated food allergy, while offering non-inferior protection against pertussis and an acceptable reactogenicity profile. To test this, we are conducting a two-stage randomised controlled trial (RCT) to assess clinical, immunological, and safety endpoints in Australian infants vaccinated with a mixed (wP/aP/aP) primary vaccine schedule, compared with the standard aP-only (aP/aP/aP) vaccine schedule.^12,13^ Ahead of ascertainment of the primary clinical outcome of IgE-mediated food allergy by 12 months old, here we present the immunogenicity and reactogenicity outcomes which were assessed in the first 150 infants enrolled in Perth, Western Australia (WA) at approximately 6 and 7 months old, as well as total IgE, TT, and food-antigen specific IgE responses at the same ages.

## Methods

### Study design and participants

The OPTIMUM (Optimising Immunisation Using Mixed Schedules) study is a Bayesian group-sequential, two-stage, multicentre, randomised, parallel group, double-blind controlled trial with an adaptive design. Stage one was designed to obtain detailed solicited reactogenicity data following each pertussis vaccine dose and post-priming immune response data for the first 150 enrolled infants. Stage two was designed to assess the primary endpoint of IgE-mediated food allergy, with less intensive follow-up, but with sufficient sample size to provide appropriate levels of statistical power. The analyses that we describe here involve the first six months of follow-up of the first 150 WA-born infants enrolled in stage one (Perth Children’s Hospital/Telethon Kids Institute, WA). The eligibility criteria, enrolment methods, and visit schedule are provided in the protocol and statistical analysis plan.^12,13^ Eligible participants were healthy infants aged between 6 and < 12 weeks old and born after 32 weeks’ gestation. The legally accepted representative(s) of each participating infant provided written informed consent. The trial was approved by the Child and Adolescent Health Service Ethics Committee, WA, Australia (RGS 00019).

### Randomisation and masking

Randomisation was by computer-generated allocation sequence prepared by the trial statistician, and based on randomly permuted blocks of size six, eight, or ten.^12,13^ Infants were assigned in a 1:1 ratio to receive either the intervention (pentavalent wP combination vaccine: (DT, TT, wP, *Haemophilus influenzae* type b [Hib], and hepatitis B [HepB] vaccine; DTwP-Hib-HepB, Pentabio, PT Bio Farma, Indonesia) or the comparator (hexavalent aP combination vaccine, which includes inactivated poliovirus vaccine [IPV] types 1, 2, and 3 in its formulation: DTaP-Hib-HepB-IPV, Infanrix Hexa, GlaxoSmithKline, Australia) at approximately 6 weeks old. An unblinded research nurse obtained the next sequential vaccine allocation and prepared it into a syringe which was covered with an opaque label. Following vaccination, this nurse had no involvement in subsequent study procedures. Parents of children in the stage one cohort were unblinded in May 2023, after the last child completed the study.

### Procedures

At approximately 6 weeks old (“Day 0”), an intramuscular dose (0.5 mL) of either a World Health Organization (WHO)-prequalified pentavalent wP combination vaccine or a hexavalent aP combination vaccine was administered into the anterolateral aspect of the right thigh. At approximately 4 and 6 months old, all participants received the standard (6-in-1) aP combination vaccine (as routinely recommended by the Australian immunisation schedule) at the same injection site (anterolateral aspect of the right thigh). The 13-valent pneumococcal conjugate vaccine (13vPCV) and monovalent rotavirus vaccine (RV1) were co-administered at approximately 6 weeks and 4 months old per Australian recommendations. National guidelines also recommend all children receive at least three doses of IPV as part of their childhood schedule; to ensure this was achieved while preserving blinding, children assigned to both the mixed and the aP-only schedule received a dose of DTaP-IPV (Infanrix-IPV, GlaxoSmithKline, Australia) at 18 months old.^14^ A prophylactic dose of paracetamol (15 mg/Kg) was administered immediately before the 6-week doses per Australian guidelines for wP vaccines.^15^ Two additional doses 6-hour apart were recommended, but not observed by the researchers.

Solicited systemic and local adverse reactions following pertussis primary vaccinations were ascertained once a day for 7 days and recorded by the participant’s parent in a diary card. These were fever (axillary temperature ≥ 38° C), irritability, restlessness, vomiting, diarrhoea, anorexia, drowsiness, as well as erythema (redness), swelling, induration (hardness) and pain at the injection-site. Serious adverse events (SAEs) were defined as any adverse event/adverse reaction that resulted in death, was life-threatening, required hospitalisation or prolongation of existing hospitalisation, resulted in persistent or significant disability or incapacity, or was a congenital anomaly, or birth defect.^16^ The total number of SAEs and adverse events of special interest (breakthrough pertussis infections and hypotonic hyporesponsive episodes) occurring within the first 6 months of follow-up were reported. SAEs and unsolicited adverse events by treatment arm will be reported at the end of the trial. To ascertain overall satisfaction of the vaccines administered, parents were asked about their agreement with the statement: “I would be willing to vaccinate another child with the same combination of vaccines administered;” responses were recorded in diary cards on Day 6 using a 5-point Likert scale, from “strongly disagree” to “strongly agree”.

Peripheral blood samples were obtained immediately before the 6-month aP dose, and approximately one month (21 to 35 days) afterwards. Serum IgG responses to diphtheria, tetanus, and pertussis antigens were measured using a multiplex fluorescent bead-based immunoassay.^17^ The threshold for seropositivity was defined as concentrations ≥ 5 IU/mL (5000 mIU/mL) for pertussis antigens (i.e., PT, filamentous haemagglutinin [FHA], pertactin [PRN], and fimbriae type 2 and 3 [FIM2/3]).^18^ Long-term seroprotective concentrations for TT and DT were defined at ≥ 100 mIU/mL.^19,20^ Total and specific IgE to hen’s egg antigens and TT were measured in plasma samples (ImmunoCAP, Thermo Fisher Scientific) using the limits of detection (LLoD) and quantitation (LLoQ) specified by the manufacturer. The total IgE concentrations were reported in the range of 0.1 kU/L to 100 kU/L for low-range assays or 2 to 5000 kU/L if exceeded. IgE concentrations to specific antigens were reported in the range of 0.00 kU_a_/L to 100.00 kU_a_/L. The LLoD for antigen/allergen specific IgE was ≥ 0.01 kU/L; the LLoQ for total and specific IgE was ≥ 0.1 kU_(a)_/L.

### Outcomes

Vaccine immunogenicity outcomes included IgG responses to PT, FHA, PRN, tetanus, and diphtheria toxoids immediately before the 6-month aP dose and approximately one month later. Outcomes related to IgE-mediated immune responses included total IgE and specific IgE against hen’s egg antigens and TT assessed immediately before the 6-month aP dose and approximately one month later. Reactogenicity and tolerability outcomes included the occurrence of specific solicited local and systemic adverse reactions in the 7 days following each scheduled dose, and the parent-reported acceptability of the doses administered.

### Statistical analysis

The statistical analysis plan was written by the trial statistician and approved by study investigators prior to unblinding of group allocations.^13^ Intention-to-treat (ITT) and per-protocol (PP) estimands were considered for each statistical model. The ITT analysis set included all infants who received at least the first dose of wP or aP, irrespective of any subsequent deviations from the study protocol. The PP analysis set included only infants with outcomes measured without deviation from the protocol-specified vaccination and blood collection schedules. Missing outcomes were assumed to be missing-at-random.

Immunogenicity and outcomes related to IgE-mediated immune responses were summarised by the sample GMCs as well as seropositivity rate at each time-point. Each antigen/allergen type was analysed using Bayesian multivariate-normal linear regression models on the available log_10_ concentrations with an unstructured covariance matrix shared across both treatment groups. The geometric mean ratio (GMR) of the mixed schedule relative to the aP-only schedule at each time-point was estimated. Analyses were conducted separately for the intent-to-treat (ITT) and per-protocol (PP) sets. A non-inferiority margin of 2/3 on the GMR was used to assess the non-inferiority of the mixed schedule versus the aP-only schedule with regards to PT-IgG approximately one month after the 6-month aP dose per WHO guidelines.^21^ To estimate the models, IgE-specific values reported as 0.00 kU_a_/L were treated as left-censored at 0.005 kU_a_/L. All adjusted models included sex, birth order, breastfeeding status, delivery method, family history of atopic disease, and parental income as baseline covariates.

To supplement the IgG and IgE models of the raw concentrations, the event of IgG seropositivity for each vaccine antigen with respect to the specified threshold and IgE concentrations ≥ 0.01 KU/L were analysed via Bayesian logistic mixed effects models. The models were used to derive standardised differences of the mixed schedule compared to the aP-only schedule for probability of seropositivity and IgE ≥ 0.01 KU/L respectively. The reactogenicity data were summarised by the distribution of each solicited adverse event for each vaccine occasion at the study site clinic and the highest/worst reaction grade experienced. Analyses were performed in R version 4.3.1 with Bayesian models fit using Stan version 2.33.1 (via CmdStanR version 0.6.1). This trial is registered at the Australian and New Zealand Clinical Trial Registry (ACTRN12617000065392p).

### Role of the funding source

This trial is funded by the Telethon New Children’s Hospital Research Fund (stage one) and Australia’s National Health and Medicine Research Council (stage two). These funding bodies had no role in the study design, data collection, data analysis, data interpretation, or writing of this report. The conduct of this trial is overseen by an independent Data and Safety Monitoring Committee.

## Results

Between March 7, 2018 and January 13, 2022, 153 infants were screened for eligibility (Figure 1), of whom 150 were randomised to wP versus aP for their 6-week pertussis vaccine dose (75 in each group). Baseline and demographic characteristics were well balanced across mixed and aP-only schedules as described in Table 1. Except for three infants in the mixed schedule arm, infants this cohort were born from an aP-vaccinated mother in the preceding pregnancy.

**Figure 1:**
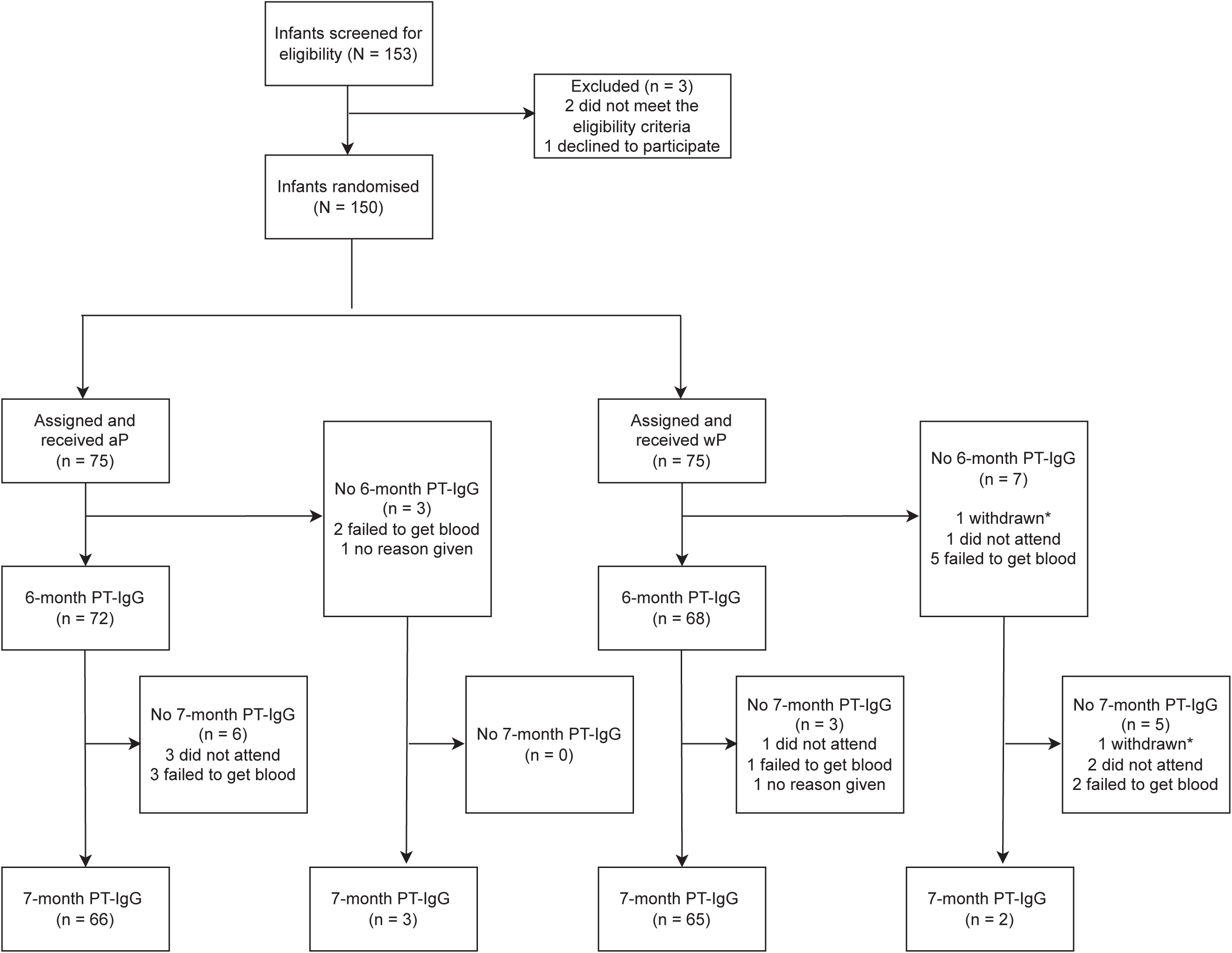
Trial profile. Legend: *Refers to the same study participant.

**Table 1.**
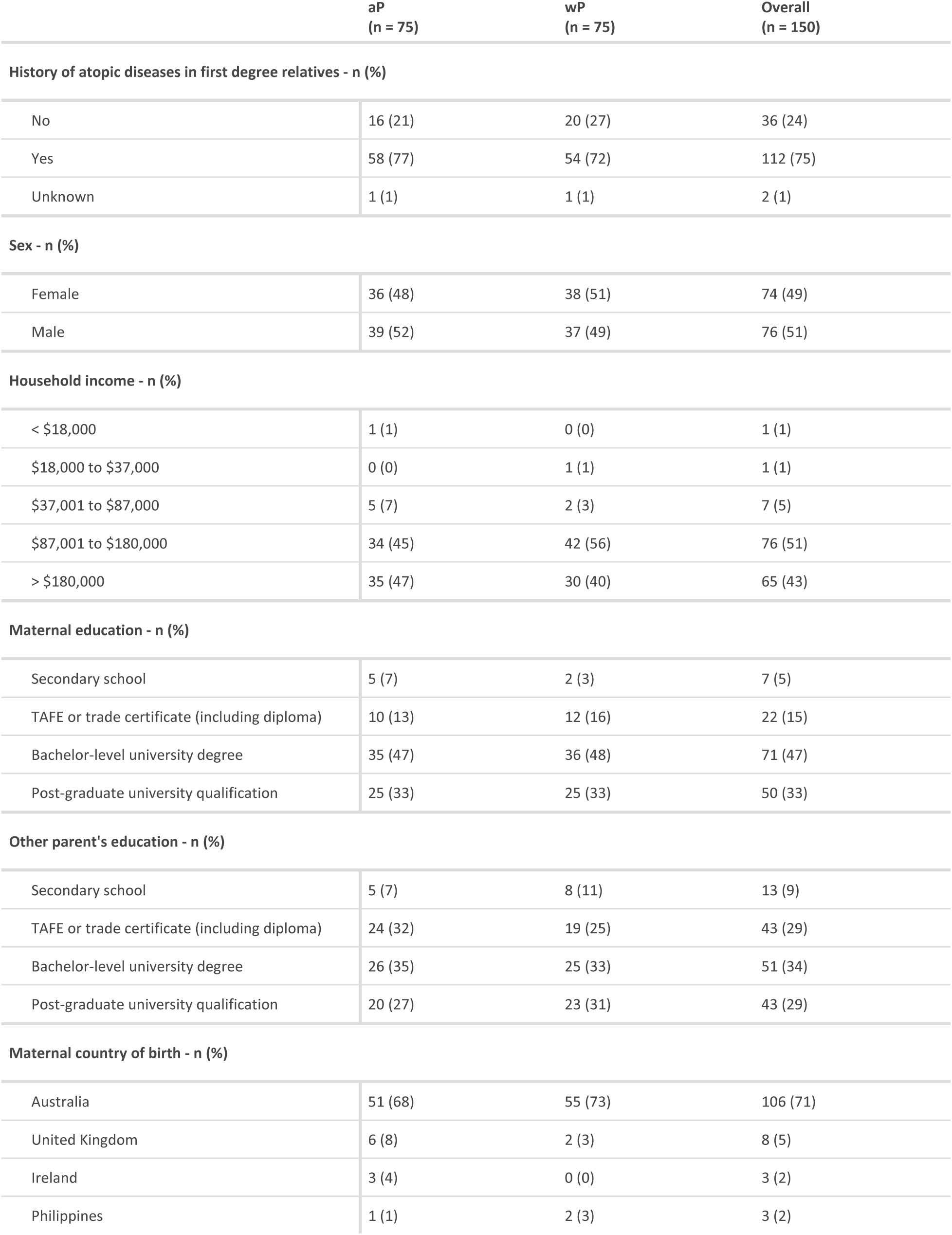
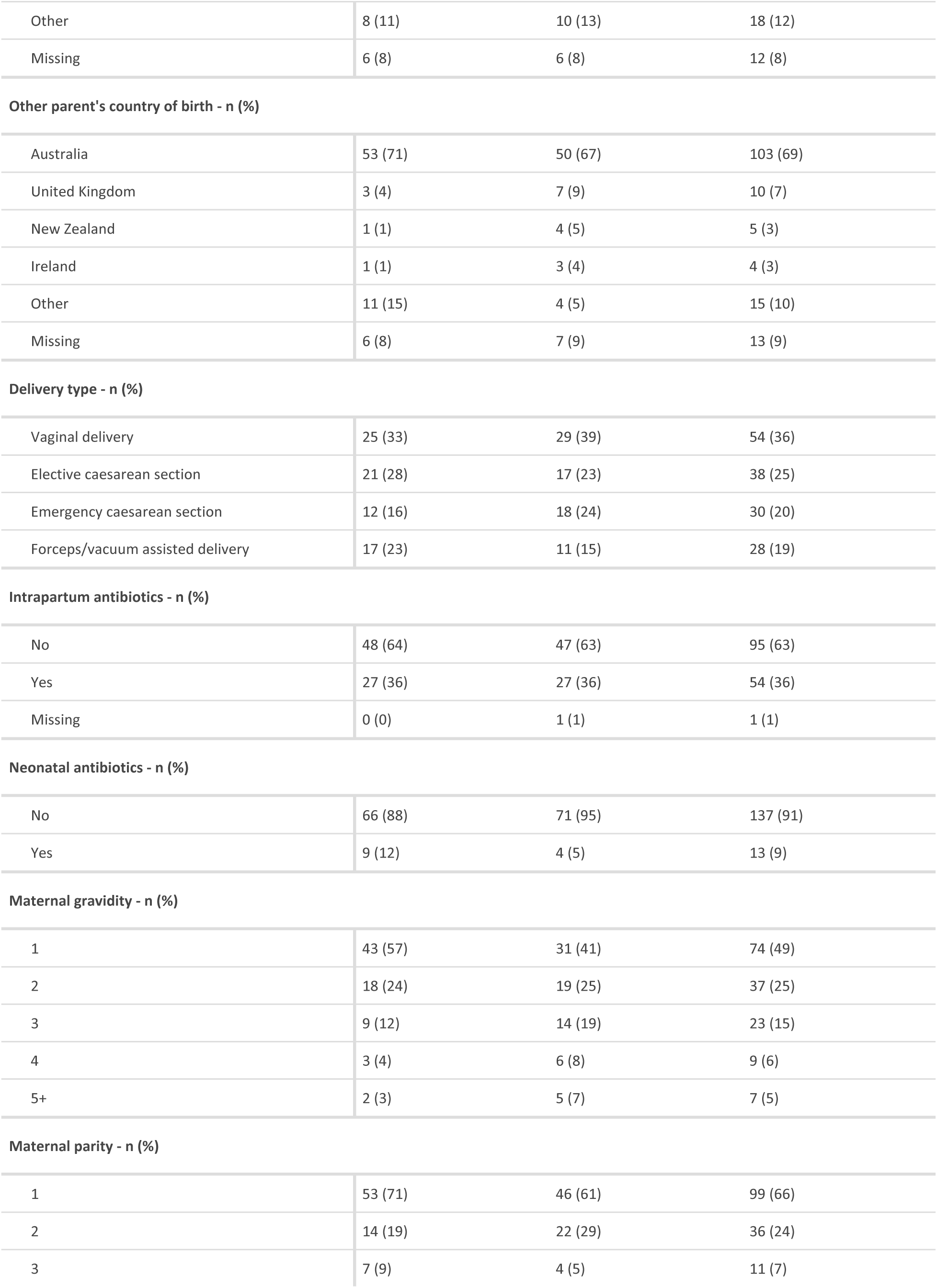
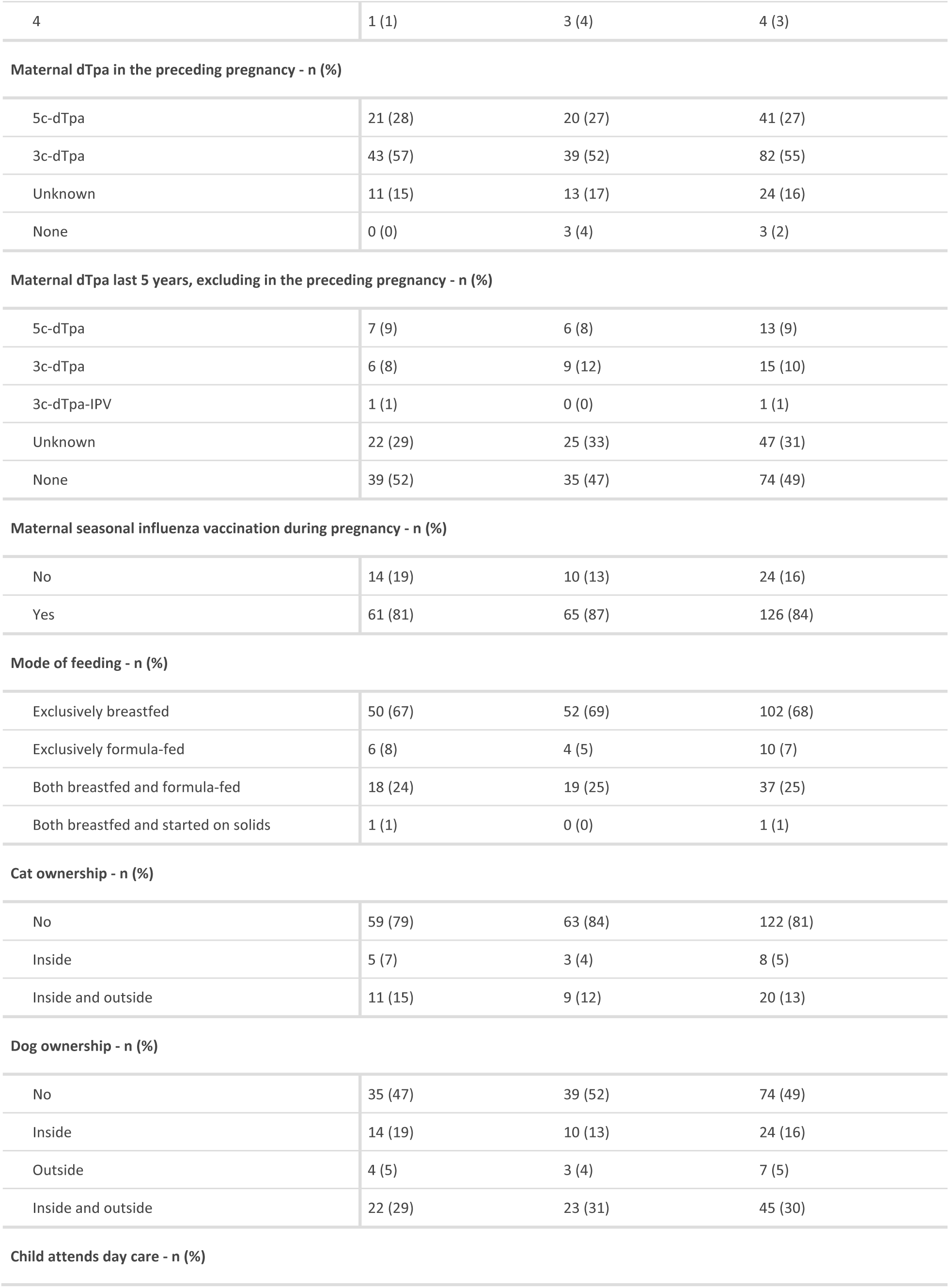
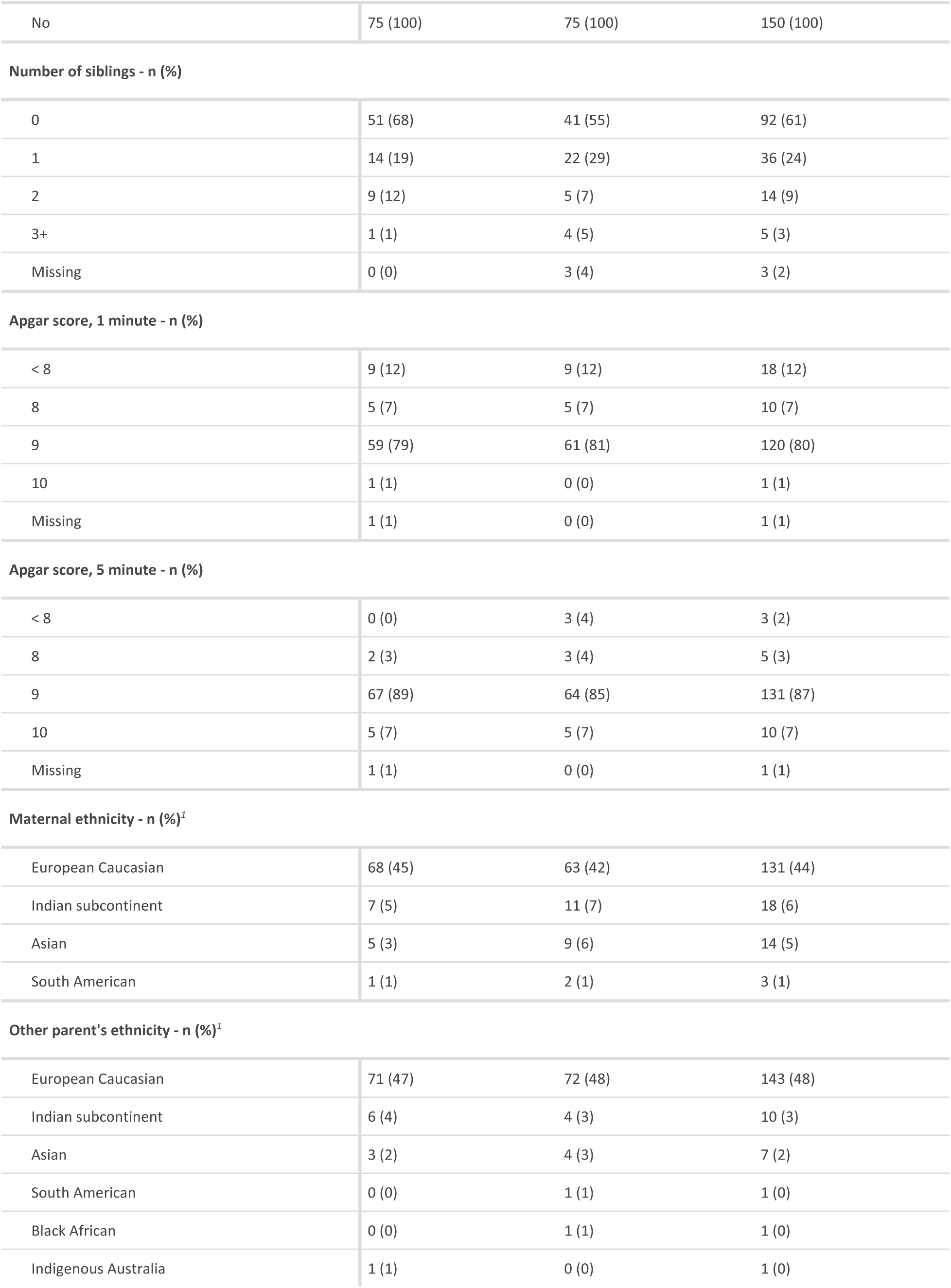
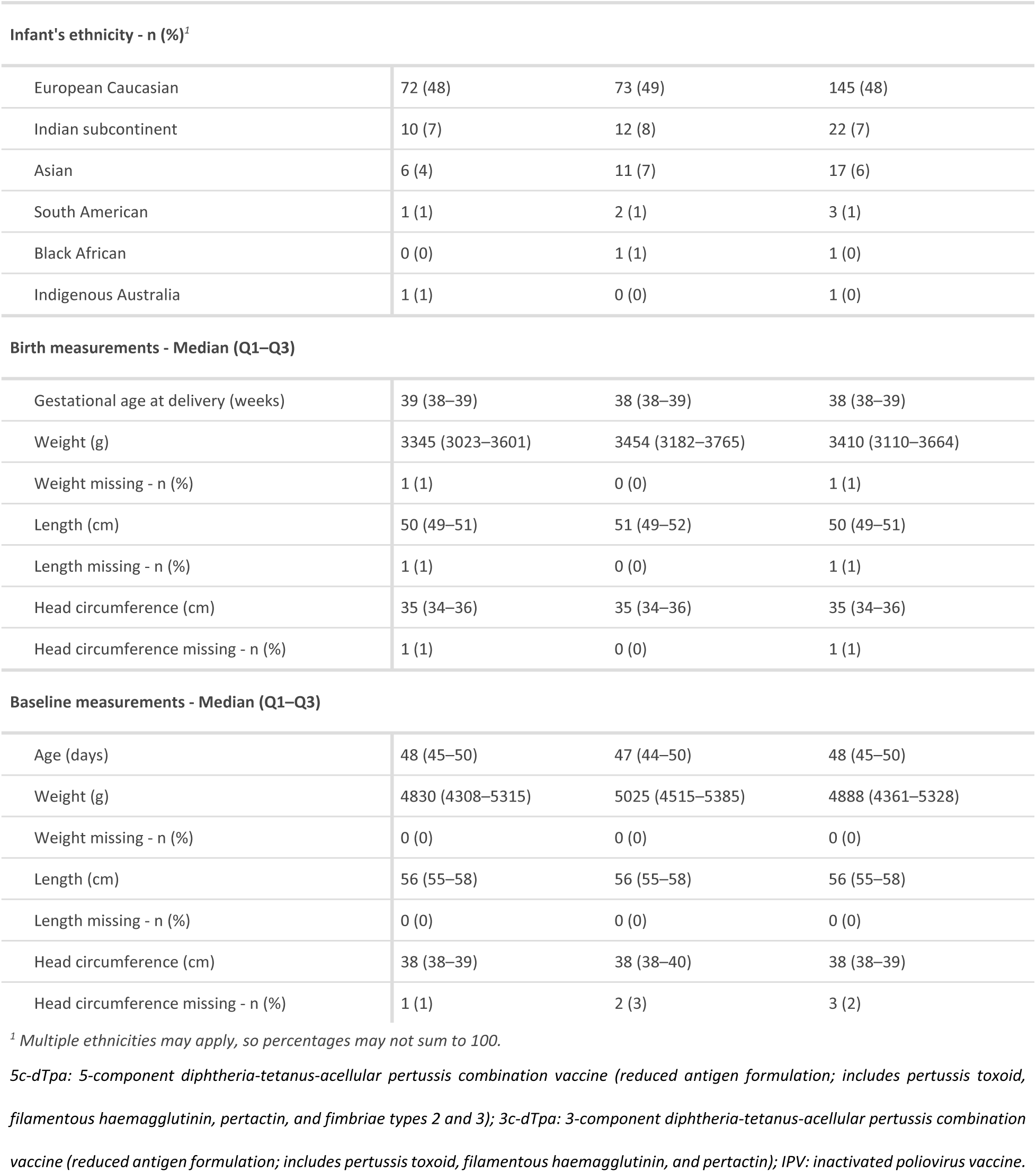
Baseline and demographic characteristics.

|  | aP<br>(n = 75) | wP<br>(n = 75) | Overall<br>(n = 150) |
| --- | --- | --- | --- |
| **History of atopic diseases in first degree relatives - n (%)** |  |  |  |
| No | 16 (21) | 20 (27) | 36 (24) |
| Yes | 58 (77) | 54 (72) | 112 (75) |
| Unknown | 1 (1) | 1 (1) | 2 (1) |
| **Sex - n (%)** |  |  |  |
| Female | 36 (48) | 38 (51) | 74 (49) |
| Male | 39 (52) | 37 (49) | 76 (51) |
| **Household income - n (%)** |  |  |  |
| < \$18,000 | 1 (1) | 0 (0) | 1 (1) |
| \$18,000 to \$37,000 | 0 (0) | 1 (1) | 1 (1) |
| \$37,001 to \$87,000 | 5 (7) | 2 (3) | 7 (5) |
| \$87,001 to \$180,000 | 34 (45) | 42 (56) | 76 (51) |
| > \$180,000 | 35 (47) | 30 (40) | 65 (43) |
| **Maternal education - n (%)** |  |  |  |
| Secondary school | 5 (7) | 2 (3) | 7 (5) |
| TAFE or trade certificate (including diploma) | 10 (13) | 12 (16) | 22 (15) |
| Bachelor-level university degree | 35 (47) | 36 (48) | 71 (47) |
| Post-graduate university qualification | 25 (33) | 25 (33) | 50 (33) |
| **Other parent's education - n (%)** |  |  |  |
| Secondary school | 5 (7) | 8 (11) | 13 (9) |
| TAFE or trade certificate (including diploma) | 24 (32) | 19 (25) | 43 (29) |
| Bachelor-level university degree | 26 (35) | 25 (33) | 51 (34) |
| Post-graduate university qualification | 20 (27) | 23 (31) | 43 (29) |
| **Maternal country of birth - n (%)** |  |  |  |
| Australia | 51 (68) | 55 (73) | 106 (71) |
| United Kingdom | 6 (8) | 2 (3) | 8 (5) |
| Ireland | 3 (4) | 0 (0) | 3 (2) |
| Philippines | 1 (1) | 2 (3) | 3 (2) |
| Other | 8 (11) | 10 (13) | 18 (12) |
| Missing | 6 (8) | 6 (8) | 12 (8) |
| **Other parent's country of birth - n (%)** |  |  |  |
| Australia | 53 (71) | 50 (67) | 103 (69) |
| United Kingdom | 3 (4) | 7 (9) | 10 (7) |
| New Zealand | 1 (1) | 4 (5) | 5 (3) |
| Ireland | 1 (1) | 3 (4) | 4 (3) |
| Other | 11 (15) | 4 (5) | 15 (10) |
| Missing | 6 (8) | 7 (9) | 13 (9) |
| **Delivery type - n (%)** |  |  |  |
| Vaginal delivery | 25 (33) | 29 (39) | 54 (36) |
| Elective caesarean section | 21 (28) | 17 (23) | 38 (25) |
| Emergency caesarean section | 12 (16) | 18 (24) | 30 (20) |
| Forceps/vacuum assisted delivery | 17 (23) | 11 (15) | 28 (19) |
| **Intrapartum antibiotics - n (%)** |  |  |  |
| No | 48 (64) | 47 (63) | 95 (63) |
| Yes | 27 (36) | 27 (36) | 54 (36) |
| Missing | 0 (0) | 1 (1) | 1 (1) |
| **Neonatal antibiotics - n (%)** |  |  |  |
| No | 66 (88) | 71 (95) | 137 (91) |
| Yes | 9 (12) | 4 (5) | 13 (9) |
| **Maternal gravidity - n (%)** |  |  |  |
| 1 | 43 (57) | 31 (41) | 74 (49) |
| 2 | 18 (24) | 19 (25) | 37 (25) |
| 3 | 9 (12) | 14 (19) | 23 (15) |
| 4 | 3 (4) | 6 (8) | 9 (6) |
| 5+ | 2 (3) | 5 (7) | 7 (5) |
| **Maternal parity - n (%)** |  |  |  |
| 1 | 53 (71) | 46 (61) | 99 (66) |
| 2 | 14 (19) | 22 (29) | 36 (24) |
| 3 | 7 (9) | 4 (5) | 11 (7) |
| 4 | 1 (1) | 3 (4) | 4 (3) |
| **Maternal dTpa in the preceding pregnancy - n (%)** |  |  |  |
| 5c-dTpa | 21 (28) | 20 (27) | 41 (27) |
| 3c-dTpa | 43 (57) | 39 (52) | 82 (55) |
| Unknown | 11 (15) | 13 (17) | 24 (16) |
| None | 0 (0) | 3 (4) | 3 (2) |
| **Maternal dTpa last 5 years, excluding in the preceding pregnancy - n (%)** |  |  |  |
| 5c-dTpa | 7 (9) | 6 (8) | 13 (9) |
| 3c-dTpa | 6 (8) | 9 (12) | 15 (10) |
| 3c-dTpa-IPV | 1 (1) | 0 (0) | 1 (1) |
| Unknown | 22 (29) | 25 (33) | 47 (31) |
| None | 39 (52) | 35 (47) | 74 (49) |
| **Maternal seasonal influenza vaccination during pregnancy - n (%)** |  |  |  |
| No | 14 (19) | 10 (13) | 24 (16) |
| Yes | 61 (81) | 65 (87) | 126 (84) |
| **Mode of feeding - n (%)** |  |  |  |
| Exclusively breastfed | 50 (67) | 52 (69) | 102 (68) |
| Exclusively formula-fed | 6 (8) | 4 (5) | 10 (7) |
| Both breastfed and formula-fed | 18 (24) | 19 (25) | 37 (25) |
| Both breastfed and started on solids | 1 (1) | 0 (0) | 1 (1) |
| **Cat ownership - n (%)** |  |  |  |
| No | 59 (79) | 63 (84) | 122 (81) |
| Inside | 5 (7) | 3 (4) | 8 (5) |
| Inside and outside | 11 (15) | 9 (12) | 20 (13) |
| **Dog ownership - n (%)** |  |  |  |
| No | 35 (47) | 39 (52) | 74 (49) |
| Inside | 14 (19) | 10 (13) | 24 (16) |
| Outside | 4 (5) | 3 (4) | 7 (5) |
| Inside and outside | 22 (29) | 23 (31) | 45 (30) |
| **Child attends day care - n (%)** |  |  |  |
| No | 75 (100) | 75 (100) | 150 (100) |
| **Number of siblings - n (%)** |  |  |  |
| 0 | 51 (68) | 41 (55) | 92 (61) |
| 1 | 14 (19) | 22 (29) | 36 (24) |
| 2 | 9 (12) | 5 (7) | 14 (9) |
| 3+ | 1 (1) | 4 (5) | 5 (3) |
| Missing | 0 (0) | 3 (4) | 3 (2) |
| **Apgar score, 1 minute - n (%)** |  |  |  |
| < 8 | 9 (12) | 9 (12) | 18 (12) |
| 8 | 5 (7) | 5 (7) | 10 (7) |
| 9 | 59 (79) | 61 (81) | 120 (80) |
| 10 | 1 (1) | 0 (0) | 1 (1) |
| Missing | 1 (1) | 0 (0) | 1 (1) |
| **Apgar score, 5 minute - n (%)** |  |  |  |
| < 8 | 0 (0) | 3 (4) | 3 (2) |
| 8 | 2 (3) | 3 (4) | 5 (3) |
| 9 | 67 (89) | 64 (85) | 131 (87) |
| 10 | 5 (7) | 5 (7) | 10 (7) |
| Missing | 1 (1) | 0 (0) | 1 (1) |
| **Maternal ethnicity - n (%)<sup>1</sup>** |  |  |  |
| European Caucasian | 68 (45) | 63 (42) | 131 (44) |
| Indian subcontinent | 7 (5) | 11 (7) | 18 (6) |
| Asian | 5 (3) | 9 (6) | 14 (5) |
| South American | 1 (1) | 2 (1) | 3 (1) |
| **Other parent's ethnicity - n (%)<sup>2</sup>** |  |  |  |
| European Caucasian | 71 (47) | 72 (48) | 143 (48) |
| Indian subcontinent | 6 (4) | 4 (3) | 10 (3) |
| Asian | 3 (2) | 4 (3) | 7 (2) |
| South American | 0 (0) | 1 (1) | 1 (0) |
| Black African | 0 (0) | 1 (1) | 1 (0) |
| Indigenous Australia | 1 (1) | 0 (0) | 1 (0) |
| **Infant's ethnicity - n (%)<sup>1</sup>** |  |  |  |
| European Caucasian | 72 (48) | 73 (49) | 145 (48) |
| Indian subcontinent | 10 (7) | 12 (8) | 22 (7) |
| Asian | 6 (4) | 11 (7) | 17 (6) |
| South American | 1 (1) | 2 (1) | 3 (1) |
| Black African | 0 (0) | 1 (1) | 1 (0) |
| Indigenous Australia | 1 (1) | 0 (0) | 1 (0) |
| **Birth measurements - Median (Q1–Q3)** |  |  |  |
| Gestational age at delivery (weeks) | 39 (38–39) | 38 (38–39) | 38 (38–39) |
| Weight (g) | 3345 (3023–3601) | 3454 (3182–3765) | 3410 (3110–3664) |
| Weight missing - n (%) | 1 (1) | 0 (0) | 1 (1) |
| Length (cm) | 50 (49–51) | 51 (49–52) | 50 (49–51) |
| Length missing - n (%) | 1 (1) | 0 (0) | 1 (1) |
| Head circumference (cm) | 35 (34–36) | 35 (34–36) | 35 (34–36) |
| Head circumference missing - n (%) | 1 (1) | 0 (0) | 1 (1) |
| **Baseline measurements - Median (Q1–Q3)** |  |  |  |
| Age (days) | 48 (45–50) | 47 (44–50) | 48 (45–50) |
| Weight (g) | 4830 (4308–5315) | 5025 (4515–5385) | 4888 (4361–5328) |
| Weight missing - n (%) | 0 (0) | 0 (0) | 0 (0) |
| Length (cm) | 56 (55–58) | 56 (55–58) | 56 (55–58) |
| Length missing - n (%) | 0 (0) | 0 (0) | 0 (0) |
| Head circumference (cm) | 38 (38–39) | 38 (38–40) | 38 (38–39) |
| Head circumference missing - n (%) | 1 (1) | 2 (3) | 3 (2) |

<sup>1</sup> Multiple ethnicities may apply, so percentages may not sum to 100.
*5c-dTpa: 5-component diphtheria-tetanus-acellular pertussis combination vaccine (reduced antigen formulation; includes pertussis toxoid, filamentous haemagglutinin, pertactin, and fimbriae types 2 and 3); 3c-dTpa: 3-component diphtheria-tetanus-acellular pertussis combination vaccine (reduced antigen formulation; includes pertussis toxoid, filamentous haemagglutinin, and pertactin); IPV: inactivated poliovirus vaccine.*

All available IgG responses are summarised in Figure 2 and in the supplementary document; per protocol analyses are included in the supplementary document.

**Figure 2.**
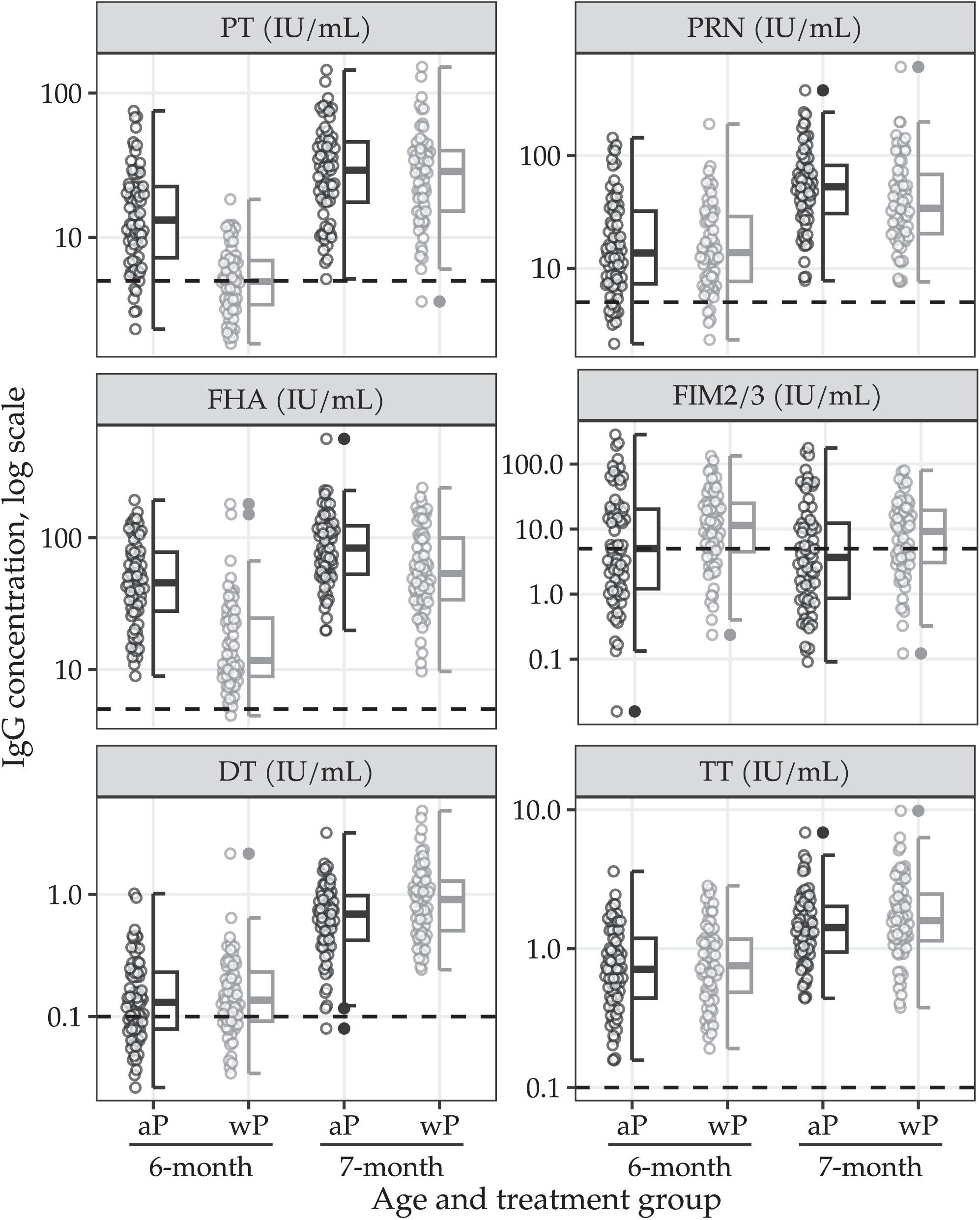
IgG concentration boxplots by antigen type, age, and assigned treatment, pertussis and other antigens. Horizontal dotted line indicates antigen-specific seropositive threshold. *Legend: PT, pertussis toxin; PRN, pertactin, FIM2/3, fimbriae 2/3; FHA, filamentous hemagglutinin DT, diphtheria toxin; TT, tetanus toxoid*.

**Figure 3.**
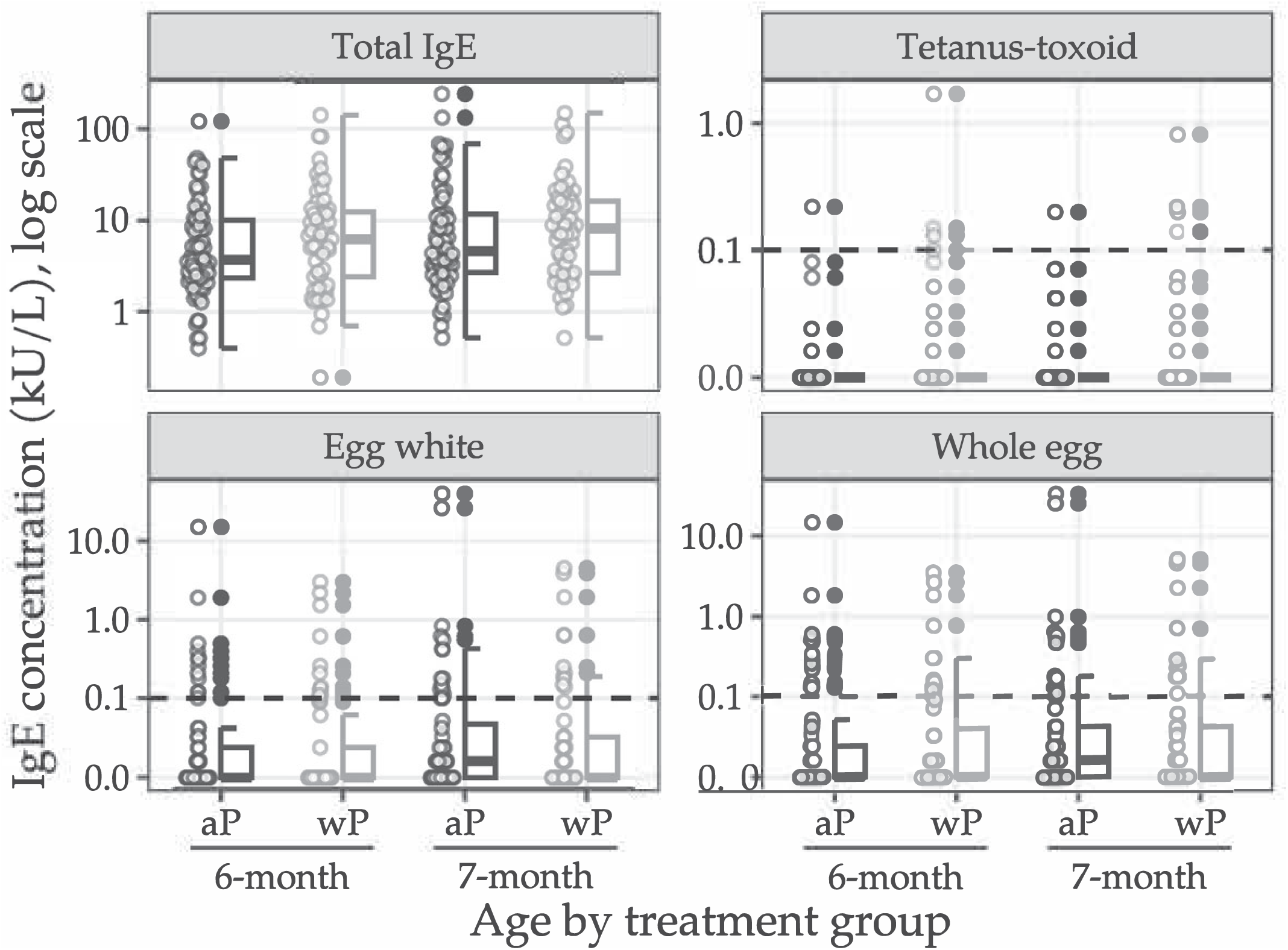
IgE concentrations by allergen type, age, and assigned schedule.

At 6 months old, 72 of 75 infants (96%) in the aP-only schedule group and 68 of 75 infants (91%) in the mixed schedule group had immunogenicity outcome data available (Figure 1). At 7 months old, 69 of 75 infants (92%) in the aP-only schedule group and 67 of 75 infants (89%) in the mixed schedule group had immunogenicity outcome data available (Figure 1).

At 7 months old, 68 of 69 infants in the aP-only group (99%) and 66 of 67 infants in the mixed schedule group (99%) had PT-, FHA-, PRN-, TT-, and DT-IgG concentrations above the specified seropositivity/seroprotective thresholds (see supplementary document). In the mixed schedule group, infants had PT-IgG GMC of 4·9 IU/mL at approximately 6 months old which increased to 26·4 IU/mL at approximately 7 months old. In the aP-only schedule group, infants had PT-IgG GMC of 13·7 IU/mL at approximately 6 months old which increased to 27·7 IU/mL at approximately 7 months old. The posterior median GMR for PT-IgG (mixed schedule/aP-only schedule) at 7 months old was 0·98 (95% Bayesian credible interval [CrI] 0·77 to 1·26; probability (GMR > 2/3) > 0·99). A decrease in IgG GMCs for FIM 2/3 and an increase in IgG GMCs for FHA, PRN, DT, and TT were observed at 7 months compared to 6 months old for both study groups (see supplementary document). The remaining IgG GMR posterior summaries are included in Table 2.

**Table 2.** IgG GMR posterior summaries (mixed schedule group/aP-only group) and non-inferiority probability, diphtheria, tetanus, and pertussis antigens.

|  | *Mean ± std* | *Median* | *95% CrI* | *Pr (> 1)* | *Pr (> 2/3)* |
| --- | --- | --- | --- | --- | --- |
| **Diphtheria toxoid** |  |  |  |  |  |
| 6-month | 1.11 ± 0.14 | 1.10 | (0.85, 1.42) | 0.77 | > 0.99 |
| 7-month | 1.36 ± 0.16 | 1.35 | (1.07, 1.70) | 0.99 | > 0.99 |
| 7/6-month (GMFR) | 1.24 ± 0.14 | 1.23 | (0.97, 1.55) | 0.96 | — |
| **Filamentous haemagglutinin** |  |  |  |  |  |
| 6-month | 0.36 ± 0.05 | 0.36 | (0.28, 0.46) | 0.00 | 0.00 |
| 7-month | 0.70 ± 0.08 | 0.69 | (0.56, 0.87) | 0.00 | 0.67 |
| 7/6-month (GMFR) | 1.97 ± 0.24 | 1.96 | (1.55, 2.47) | > 0.99 | — |
| **Fimbriae 2/3** |  |  |  |  |  |
| 6-month | 1.96 ± 0.59 | 1.88 | (1.04, 3.34) | 0.98 | > 0.99 |
| 7-month | 1.85 ± 0.53 | 1.78 | (1.01, 3.09) | 0.98 | > 0.99 |
| 7/6-month (GMFR) | 0.95 ± 0.07 | 0.95 | (0.82, 1.09) | 0.23 | — |
| **Pertactin** |  |  |  |  |  |
| 6-month | 1.03 ± 0.17 | 1.01 | (0.74, 1.40) | 0.53 | > 0.99 |
| 7-month | 0.80 ± 0.12 | 0.79 | (0.59, 1.07) | 0.06 | 0.88 |
| 7/6-month (GMFR) | 0.79 ± 0.13 | 0.78 | (0.57, 1.06) | 0.06 | — |
| **Pertussis toxin** |  |  |  |  |  |
| 6-month | 0.37 ± 0.04 | 0.37 | (0.30, 0.46) | 0.00 | 0.00 |
| 7-month | 0.99 ± 0.13 | 0.98 | (0.77, 1.26) | 0.43 | > 0.99 |
| 7/6-month (GMFR) | 2.64 ± 0.27 | 2.63 | (2.15, 3.21) | > 0.99 | — |
| **Tetanus toxoid** |  |  |  |  |  |
| 6-month | 1.16 ± 0.14 | 1.15 | (0.92, 1.45) | 0.89 | > 0.99 |
| 7-month | 1.16 ± 0.12 | 1.15 | (0.93, 1.42) | 0.91 | > 0.99 |
| 7/6-month (GMFR) | 1.01 ± 0.10 | 1.00 | (0.83, 1.21) | 0.50 | — |

*GMR, geometric mean ratio; std, standard deviation; CrI: credible interval; Pr, probability; GMFR, geometric mean fold rise*

Total IgE and IgE responses to hen’s egg and TT antigens are summarised in Table 3, Figure3, and in the supplementary document.

**Table 3.** Sample IgE concentration summaries (kU_[a]_/L) by allergen type and assigned vaccination schedule.

|  | *aP* |  |  |  |  |  | *wP* |  |  |  |  |  |
| --- | --- | --- | --- | --- | --- | --- | --- | --- | --- | --- | --- | --- |
|  | n | GMC | Q1–Q3 | Min–Max | ≥0.01 | ≥0.1 | n | GMC | Q1–Q3 | Min–Max | ≥0.01 | ≥0.1 |
| **Total IgE** |  |  |  |  |  |  |  |  |  |  |  |  |
| 6-month | 67 | 4.76 | 2.34–10.11 | 0.40–121.43 | 100% | 100% | 61 | 6.11 | 2.40–12.43 | 0.19–141.38 | 100% | 100% |
| 7-month | 66 | 6.36 | 2.69–11.72 | 0.52–241.99 | 100% | 100% | 57 | 7.71 | 2.66–16.30 | 0.52–148.70 | 100% | 100% |
| **Tetanus toxoid IgE** |  |  |  |  |  |  |  |  |  |  |  |  |
| 6-month | 67 | 0.00 | 0.00–0.00 | 0.00–0.22 | 12% | 1% | 61 | 0.00 | 0.00–0.00 | 0.00–1.71 | 21% | 7% |
| 7-month | 66 | 0.00 | 0.00–0.00 | 0.00–0.20 | 21% | 3% | 57 | 0.00 | 0.00–0.00 | 0.00–0.82 | 21% | 7% |
| **Egg white IgE** |  |  |  |  |  |  |  |  |  |  |  |  |
| 6-month | 67 | 0.00 | 0.00–0.02 | 0.00–14.91 | 36% | 18% | 61 | 0.00 | 0.00–0.02 | 0.00–3.00 | 26% | 20% |
| 7-month | 67 | 0.00 | 0.00–0.04 | 0.00–39.50 | 54% | 24% | 57 | 0.00 | 0.00–0.03 | 0.00–4.51 | 42% | 19% |
| **Whole egg IgE** |  |  |  |  |  |  |  |  |  |  |  |  |
| 6-month | 67 | 0.00 | 0.00–0.02 | 0.00–14.85 | 40% | 18% | 60 | 0.00 | 0.00–0.04 | 0.00–3.51 | 42% | 22% |
| 7-month | 67 | 0.00 | 0.00–0.04 | 0.00–33.70 | 52% | 22% | 57 | 0.00 | 0.00–0.04 | 0.00–5.16 | 40% | 21% |

*GMC, geometric mean concentrations. Q1–Q3: first and third quartile. Min–Max, range: minimum and maximum. Values of 0.00 set to 0.005 for* *calculation of sample GMC.*

At approximately 7 months old, total IgE levels ≥ 0.1 kU/L were observed in all infants assigned to the mixed schedule (GMC = 7.71 kU/L) and to the aP-only schedule (GMC = 6·36 kU/L; Table 3). The posterior median GMR for total IgE (mixed schedule /aP-only schedule) was 1·14 (95% CrI 0·75 to 1·72; probability (GMR < 1) 0·27). The estimated posterior risk and risk difference between the mixed schedule group and the aP-only group for TT, egg white, and whole egg IgE are presented in Table 4 and in the supplementary document.

**Table 4.** Posterior risk and risk difference for tetanus toxoid and hen’s egg IgE quantitation by age and pertussis schedule.

|  | *Marginal Pr (IgE ≥ 0.01), Median (95% CrI)* |  |  | *Odds ratios* |  |  |
| --- | --- | --- | --- | --- | --- | --- |
|  | *aP* | *wP* | *wP-aP* | *Pr (wP-aP <0)* | *Conditional* | *Marginal* |
| **Tetanus toxoid IgE** |  |  |  |  |  |  |
| Unadjusted |  |  |  |  |  |  |
| 6-month | 0.15 (0.07,0.27) | 0.21 (0.11, 0.34) | 0.05 (-0.05,0.17) | 0.17 | 2.69 (0.35,28.27) | 1.40 (0.71, 3.39) |
| 7-month | 0.22 (0.12, 0.35) | 0.21 (0.11, 0.34) | -0.01 (-0.14, 0.13) | 0.53 | 0.91 (0.07, 9.44) | 0.97 (0.43, 2.14) |
| Adjusted |  |  |  |  |  |  |
| 6-month | 0.16 (0.08, 0.27) | 0.21 (0.11, 0.33) | 0.04 (-0.05, 0.15) | 0.20 | 2.61 (0.27, 35.40) | 1.33 (0.67, 3.11) |
| 7-month | 0.22 (0.12, 0.34) | 0.22 (0.12, 0.34) | 0.00 (-0.12, 0.12) | 0.52 | 0.95 (0.06, 13.91) | 0.98 (0.43, 2.26) |
| **Egg white IgE** |  |  |  |  |  |  |
| Unadjusted |  |  |  |  |  |  |
| 6-month | 0.36 (0.24, 0.49) | 0.27 (0.16, 0.40) | -0.09 (-0.23, 0.05) | 0.90 | 0.37 (0.06, 1.69) | 0.66 (0.33, 1.23) |
| 7-month | 0.51 (0.38, 0.65) | 0.44 (0.30, 0.59) | -0.07 (-0.23, 0.09) | 0.80 | 0.50 (0.09, 2.60) | 0.76 (0.38, 1.44) |
| Adjusted |  |  |  |  |  |  |
| 6-month | 0.35 (0.24, 0.48) | 0.28 (0.17, 0.41) | -0.07 (-0.20, 0.05) | 0.86 | 0.36 (0.04, 2.35) | 0.72 (0.35, 1.31) |
| 7-month | 0.49 (0.37, 0.62) | 0.46 (0.33, 0.59) | -0.03 (-0.19, 0.11) | 0.68 | 0.63 (0.07, 5.67) | 0.86 (0.43, 1.67) |
| **Whole egg IgE** |  |  |  |  |  |  |
| Unadjusted |  |  |  |  |  |  |
| 6-month | 0.41 (0.29, 0.55) | 0.40 (0.27, 0.54) | -0.01 (-0.15, 0.13) | 0.57 | 0.85 (0.14, 4.90) | 0.95 (0.52, 1.71) |
| 7-month | 0.51 (0.37, 0.64) | 0.42 (0.28, 0.56) | -0.09 (-0.24, 0.07) | 0.87 | 0.35 (0.05, 2.26) | 0.71 (0.36, 1.32) |
| Adjusted |  |  |  |  |  |  |
| 6-month | 0.41 (0.30, 0.54) | 0.41 (0.28, 0.53) | -0.01 (-0.13, 0.12) | 0.55 | 0.88 (0.11, 6.97) | 0.96 (0.55, 1.72) |
| 7-month | 0.49 (0.37, 0.62) | 0.41 (0.28, 0.53) | -0.06 (-0.21, 0.08) | 0.81 | 0.36 (0.03, 3.55) | 0.75 (0.38, 1.43) |

*Pr*, probability, *CrI*, credible interval; *aP*, acellular pertussis vaccine; *wP*, whole-cell pertussis vaccine. All adjusted models included sex, birth order, breastfeeding status, delivery method, family history of atopic disease, and parental income as baseline covariates.

The distribution of daily grades for solicited local and systemic adverse reactions following each vaccine occasion are presented in Figures 4, 5, and 6 and in the supplementary document. After the 6-week doses, 74 of 75 (99%) diary cards were returned in the mixed schedule group and 72 of 75 (96%) in the aP-only schedule group. Paracetamol use after the 6-week vaccine doses on more than one occasion was recorded among 59 of 75 (79%) wP and 52 of 75 (69%) aP recipients. A first dose of wP was more frequently followed by mild-to-moderate solicited local and systemic adverse reactions than aP (see supplementary document).

**Figure 4.**
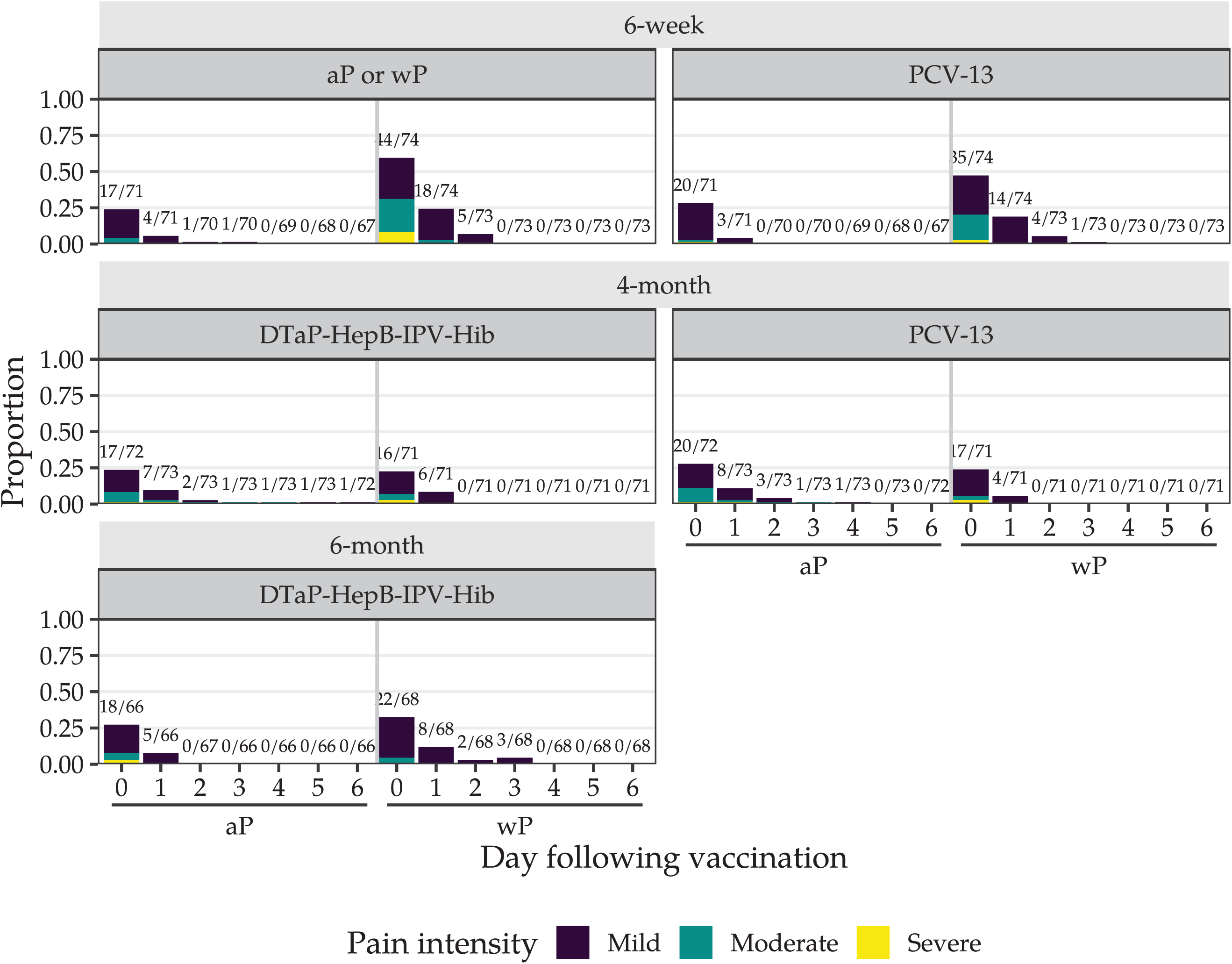
Maximum injection site reaction size in the 7 days after the 6-week vaccine doses.

Self-limited grade 3 (severe) erythema and swelling were reported at the pertussis vaccine (1 of 74; 1%) and 13vPCV (1 of 74; 1%) injection-sites after a 6-week wP dose; none were reported after the 6-week aP dose (Figure 5 and supplementary document). No reports of severe erythema, swelling, or induration at the pertussis vaccine or 13vPCV injection-sites were recorded in either study group after the 4-month or 6-month vaccine doses. Severe pain was recorded at the pertussis vaccine injection-site after a 6-week wP dose (6 of 74; 8%), but not after a 6-week aP dose (0 of 72).

**Figure 5.**
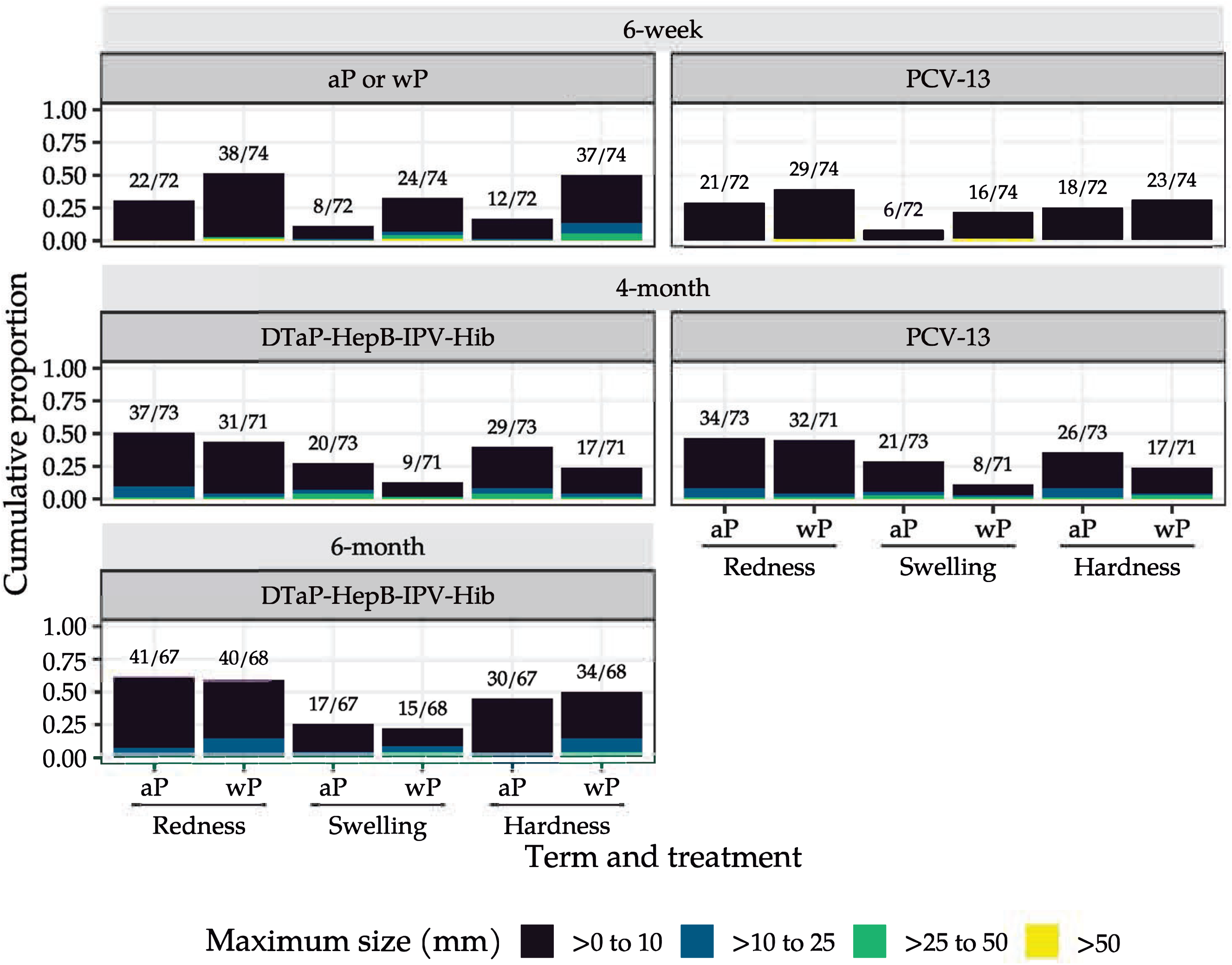
Injection site pain in the 7 days after the 6-week vaccine doses.

**Figure 6.**
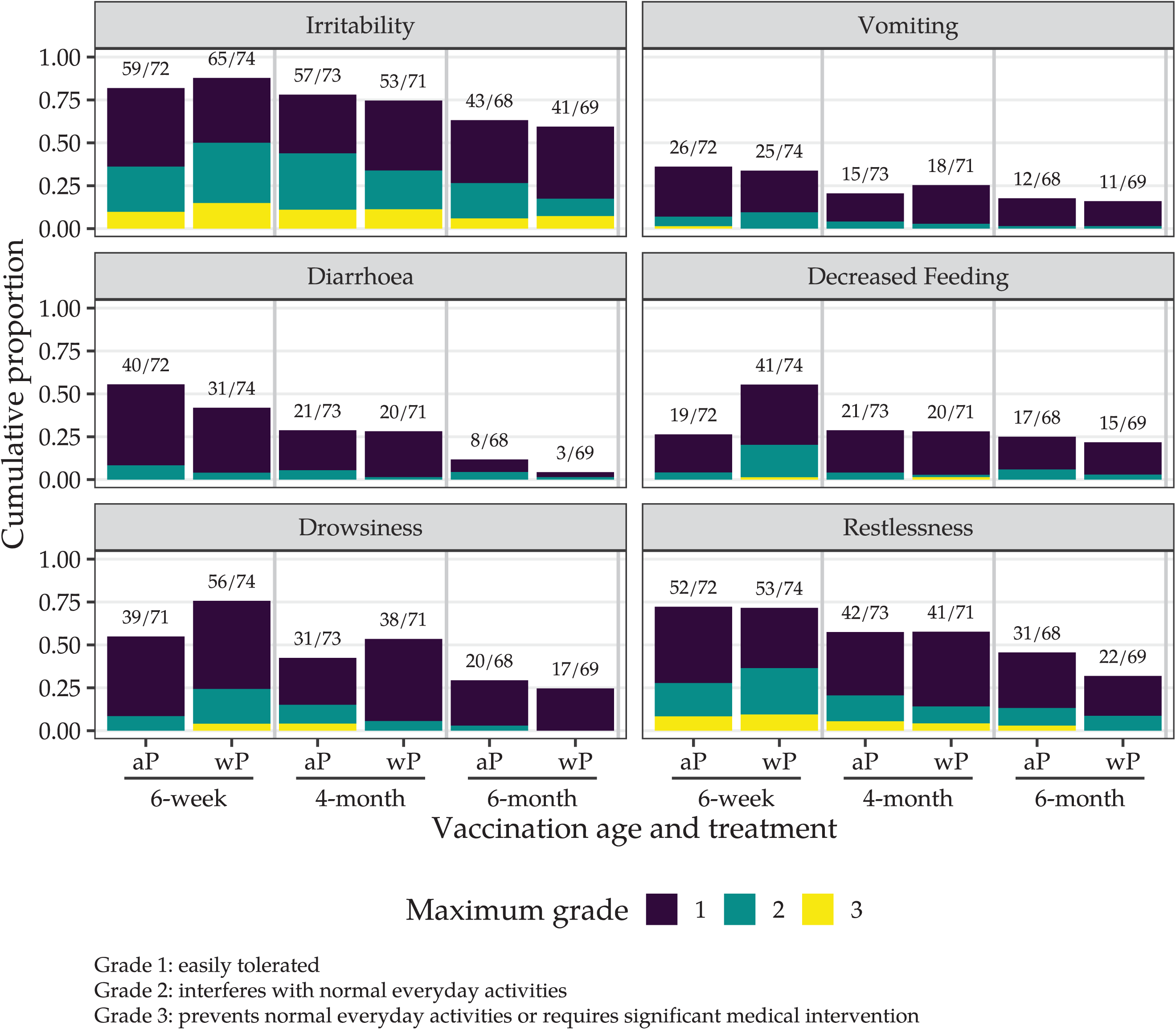
Highest intensity grade for systemic reactions after the 6-week vaccine doses.

Fever ≥ 38.0° C was reported in 0 of 73 infants vaccinated with wP and 1 of 70 (1%) infants vaccinated with aP at 6 weeks old, in 3 of 70 infants (4%) in the mixed schedule group and in 4 of 72 (6%) infants in the aP-only schedule group at 4 months old, and in 1 of 68 infants (1%) in each study group at 6 months old.

The proportion of infants who experienced one or more severe (grade 3) solicited systemic adverse reactions was higher among those who received wP (14/74; 19%) than aP (8/72; 11%) at 6 weeks old, but similar after the 4-month (mixed schedule 9 of 71; 13% versus aP-only schedule 12 of 73; 16%) and 6-month aP doses (mixed schedule 5 of 69; 7% versus aP-only schedule 5 of 68; 7%). Severe irritability was reported in 11 of 74 (15%) wP versus 7 of 72 (10%) aP recipients at 6 weeks old, in 8 of 71 (11%) in the mixed schedule group and 8 of 73 (11%) in the aP-only schedule group at 4 months old, and in 5 of 69 (7%) in the mixed schedule group versus 4 of 68 (6%) in the aP-only schedule group at 6 months old.

On Day 6 post-vaccination at 6 weeks’ old, 71 of 73 (97%) parents of wP recipients and 69 of 72 (96%) parents of aP recipients either agreed or strongly agreed that they would be happy for their child to receive the same vaccine combination.

There were 7 SAEs among 5 participants within the first 6 months of follow-up; on blinded assessment, none were assessed to be related to the study vaccines. No hypotonic hyporesponsive episodes or breakthrough pertussis infections occurred in either group.

## Discussion

We report the results of a randomised comparison of the immunogenicity, reactogenicity, and IgE-mediated immune responses to a mixed wP/aP primary vaccine schedule versus the standard aP-only schedule. Mixed priming (wP/aP/aP) using the WHO prequalified 5-in-1 wP vaccine (Pentabio, PT Biofarma, Indonesia) and 3c-aP vaccine (Infanrix Hexa, GlaxoSmithKline, Australia) formulation was non-inferior to the homologous aP-only schedule (aP/aP/aP) with respect to anti-PT antibody titres at 6 and 7 months old. While almost all (99%) serum responses to DT, TT, FHA, and PRN in the two treatment groups met their specified seropositivity/seroprotective thresholds at 7 months old, PT and FHA IgG concentrations immediately before the 6-month aP dose were lower in the mixed schedule group compared to the aP-only group. Given the lower content of PT and FHA in wP versus aP-based vaccines, this finding was not unexpected; by one month after the 6-month aP dose, PT concentrations were similar, but concentrations of FHA remained lower in the mixed schedule compared to the aP-only schedule group.

Previous studies have found concentrations of antibodies to aP vaccine antigens were either similar or lower in infants primed with wP than in those primed with aP.^22^ In addition, it has been speculated that maternal vaccination might cause greater interference of the immunogenicity of wP than aP vaccines in early infancy.^22^ In our trial, almost all mothers had received aP in pregnancy, so their infants are likely to have had high levels of maternally-derived antibodies targeting aP vaccine antigens at the time of receipt of the 6-week vaccine doses.

A dose of wP at approximately 6 weeks old was more reactogenic than a dose of aP, with a higher frequency of solicited local and systemic adverse reactions which were mostly mild-to-moderate in severity. While after the 6-week doses severe local adverse reactions were seldom reported and only described at the wP and 13vPCV injection sites, fever ≥ 38.0°C was only described in the aP-vaccine study group. The proportion of parents of wP-vaccinated infants who reported acceptability of the vaccination was very high and similar to that of parents of aP-vaccinated infants. The young age of administration of first dose of the vaccines and the routine use of prophylactic paracetamol may have attenuated the intensity of solicited reactions and possibly enhanced parental acceptance of wP.^23^ The reactogenicity data were presented quarterly to an independent data safety and monitoring committee which supported continued enrolment.

A previous RCT suggested that therapeutic paracetamol administered within 48 hours post DTwP vaccination does not interfere with IgG immune responses elicited by the vaccine antigens in children receiving a homologous 2, 4, 6, and 18-month schedule.^24^ More recently, a clinical trial reported that prophylaxis with oral paracetamol had no effect on immunogenicity of a combination aP vaccine, co-administered with 7v-PCV and a recombinant multicomponent meningococcal B vaccine at 2, 3, and 4 months old.^25^ By contrast, in infants receiving a combination aP vaccine co-administered with the 10-valent pneumococcal non-typeable *Haemophilus influenzae* protein D-conjugate vaccine (PhiD-CV) at 3, 4, and 5-months old, prophylactic rectal paracetamol was associated with reduced IgG responses to aP antigens and pneumococcal vaccine polysaccharides.^26^ While the clinical significance of these findings is uncertain, a follow-up study suggested that paracetamol prophylaxis had no impact on the induction of PhiD-CV serotype-specific immunological memory or pneumococcal nasopharyngeal carriage as measured at 4 years old.^27^

Owing to evidence of improved long term pertussis protection among wP vaccine recipients, the WHO recommends that countries using wP should only changeover to less reactogenic aP-only schedules where it is financially and programmatically feasible to provide frequent aP boosters, including in pregnancy.^28^ For countries which have already transitioned to an aP-only schedules, mixed schedules may provide better long-lived protection against pertussis but confirmation of this would require the follow-up of many thousands of infants over many years.

Examining the immune responses driven by wP vaccine formulations is complex. The exact concentrations of pertussis antigens within wP varies across formulations. Previous clinical trials reported differences in immune responses between wP and aP-based vaccines administered in homologous priming schedules and across various specific formulations of the two vaccine types. Antibody responses to aP-specific antigens are generally higher among aP-vaccinated infants than wP-vaccinated infants.^29–31^ However, greater vaccine effectiveness has been documented among school-aged children and adolescents who had received wP versus aP as their first dose.^4^ Pertussis-specific T-cell memory responses may be important in long-term protection after wP combination vaccine and will be evaluated in these infants.

Observational studies suggest that laboratory-confirmed pertussis may be less likely among wP-primed versus aP-primed children, and among those receiving mixed schedules in which the first dose was wP versus aP.^3,32^ A case-control study found that pertussis disease was less common among children receiving a mixed schedule of wP/3c-aP versus 3c-aP only. No evidence was found of a difference in the risk of pertussis among those vaccinated with a mixed wP/5c-aP schedule versus those vaccinated with a with a 5c-aP formulation, although long-term differences cannot be excluded.^33^ Another case-control study found that compared to a 5c-aP-only primary series, vaccination with one or more primary doses of a low-efficacy wP vaccine formulation was associated with a lower risk of pertussis disease more than a decade after priming.^32^ None of the cited case-control analyses provided further details regarding the nature of the first pertussis vaccine dose in the mixed schedules examined.

WHO-prequalified 5-in-1 wP vaccine formulations have been successfully introduced across 77 lower-income countries supported by Gavi. While earlier concerns about safety led to the discontinuation of wP in Australia and other high-income countries, a 2021 meta-analysis of 15 RCTs (38,072 infants) found that any risk difference for SAEs in infants primed with wP versus aP is likely to be small, ranging from three fewer to two more events per 1,000 children.^34^ Similar to our trial, the meta-analysis defined SAEs as event that resulted in death, were life-threatening, required hospitalisation or prolongation of existing hospitalisation, or resulted in persistent or significant disability or incapacity.^34^ Our data are therefore consistent with the excellent safety record of wP previously highlighted by the WHO, and support the ongoing use of wP.

We have previously observed that Australian children with IgE-mediated food allergy were less likely than contemporaneous controls to have received one or more doses of wP in infancy.^5^ We hypothesised that compared to a standard aP-only schedule, a mixed vaccine schedule comprising an initial dose of wP could help promote the normal transition from a Th_2_-skewed to a balanced Th_1_/Th_17_/Th_2_ immunophenotype in early infancy, and thereby protect against IgE-mediated food allergy. Having previously noted that wP-primed infants were less likely than aP-primed infants to develop boosted vaccine-associated IgE responses,^9^ we sought to confirm this observation by testing for total, TT, and egg-specific IgE at 6 and 7 months. While the production of specific IgE against DTaP antigens is known to only occur in a subset of vaccinated infants, children, or adults, the low TT-IgE concentrations observed in our study are not easily explained. The detection of total and specific IgE were conducted using an autoanalyser IgE-based assay and therefore interference by anti-allergen specific IgG is unlikely.^35^ However, except for analyses carried out in Australia,^9^ Belgium,^7^ and the US,^36^ most paediatric TT-specific IgE response data reported in the literature have been measured using RAST or other serological methods.^8,37^ Thus, apart from the first group of cited studies, it is not possible to directly compare our IgE results with prior cohorts.

This study represents the first prospective randomised comparison of a novel mixed wP/aP versus aP-only schedule. We used a prequalified 5-in-1 combination vaccine formulation already widely adopted and available in Gavi supported countries, and the immunogenicity of each schedule was assessed using a validated multiplex fluorescent bead-based immunoassay.

The limitations of this study include uncertain generalisability since the study population comprised a high proportion of infants of privately insured urban parents. Secondly, while our previous case-control study suggests that the potential allergy protective benefits of wP might be confined to IgE-mediated peanut or tree nuts allergy,^5^ stage one did not examine IgE responses to these antigens. In addition, we were not powered to compare schedules for important clinical outcomes, including pertussis disease. The mixed schedule was well accepted by the cohort of parents who consented to participate after being informed of the known reactogenicity profile of wP; it is unclear whether the high acceptance of wP observed is generalisable to all parents.

In conclusion, our findings give support to the acceptable immunogenicity and reactogenicity of the mixed primary schedule. These findings are relevant to countries where both wP and aP vaccines are used and support the further evaluation of the clinical effects of mixed schedules. We failed to find confirming immunological evidence of an attenuating effect of the mixed versus the aP-only schedule on TT or egg-related IgE antibodies; owing to the unclear relationship between these biomarkers and the subsequent development of IgE-mediated food allergy, a clinically important effect cannot be excluded. In stage two, the effect of the mixed versus standard schedule on the development of IgE-mediated food allergy by 12 months will be assessed in up to 2000 infants using an adaptive design with prespecified stopping rules for success or futility.

## Supporting information

CONSORT Checklist

## Data Availability

Data from the study will be available at the completion of follow-up, analysis and reporting of OPTIMUM stage two; no end date. Deidentified, individual participant data and a data dictionary defining each field in the set, will be made available to researchers who provide a methodologically sound proposal to the corresponding author (TS), subject to a signed data access agreement and any necessary ethics approvals (see ANZCTR trial registration). The study protocol and statistical analysis plan are published, other study related documentation is available on request. 

https://osf.io/4cw6g/?view_only=ccb65276705f4f8daeaeaaeb89d36e91

## Contributors

Conceptualisation: JAM, KPP, DEC, CSW, NC, PBM, PGH, PCR, and TS. Data curation: GPC and JT. Formal analysis: JT and SM. Funding acquisition: JAM, KPP, DEC, NW, MG, CSW, NC, PCR, and TS. Investigation: GPC, MJE, MOS, and SM. Methodology: MJE, JT, JAM, KPP, DEC, CSW, SM, NC, PCR, and TS. Project administration: MJE. Software: JT. Supervision: MJE, MOS, PCR, and TS. Validation: JT, SM, and MJ. Access and verification of the underlying (unblinded) data reported in the manuscript: JT and MJ. Visualisation: JT. Writing – original draft: GPC. Writing – review and editing: GP, MJE, JT, JAM, KPP, DEC, NW, MG, CSW, MOS, MJ, PGH, PCR, and TS. All authors provided their input and approved the final version of this manuscript.

## Declaration of interests

Unrelated to the work presented in this paper, DEC declared part time employment in DBV Technologies, as well as DBV Technologies stock and stock options. DEC has also served on the scientific advisory board for AllerGenis.

Unrelated to the work presented in this paper, MO is the non-remunerated Director of ASCIA, a not-for-profit company and the peak professional body for allergy/immunology specialists in Australia and New Zealand.

Unrelated to the work presented in this paper, PCR has served on pertussis vaccine scientific advisory boards for GlaxoSmithKline and Sanofi on behalf of his institution. He participated in multicentre vaccine trials of pertussis vaccines sponsored by industry, also unrelated to the work presented in this paper. He has received no personal remuneration for these activities.

All other authors report no potential conflicts.

## Acknowledgement

This trial is funded by the Telethon New Children’s Hospital Research Fund (stage one) and Australia’s National Health and Medicine Research Council (stage two). GPC was supported by a Stan Perron Post-PhD Career Launching Award, the Australian Department of Education and Training Endeavour Scholarship, as well as top-up scholarships from the Wesfarmers Centre of Vaccine and Infectious Diseases at the Telethon Kids Institute and the Forrest Research Foundation. KPP is supported by an NHMRC Investigator Grant and a Melbourne Children’s Clinician-Scientist Fellowship. SM was supported by the Australian Government Research Training Program Scholarship, the Stan Perron Excellence Award, and top-up scholarships from the UWA Safety Net, Wesfarmers Centre of Vaccine and Infectious Diseases, and the Stan Perron Foundation. TS is supported by an MRFF Investigator Award (MRF1195153). We thank the parents and infants who took part in stage one of this trial for their commitment and continuous support. We acknowledge Jane Jones and all members of the Vaccine Trials Group at the Telethon Kids Institute, who were involved in the conduct of stage one of the OPTIMUM study. We would also like to express our gratitude to Kylie Rogers (data manager) and the independent data safety monitoring committee for their valuable contributions and detailed oversight. We thank Catherine Hughes (AM), consumer representative in the OPTIMUM study Steering Committee Group for her valuable input in the design and conduct of this study

## Notes

### Clinical Trial

ACTRN12617000065392p

### Clinical Protocols

https://bmjopen.bmj.com/content/10/12/e042838

https://trialsjournal.biomedcentral.com/articles/10.1186/s13063-021-05874-6

### Author Declarations

The Child and Adolescent Health Service Ethics Committee of Western Australia, Australia gave ethical approval for this work.

