## Supplementary material for "Immunogenicity, reactogenicity, and IgE-mediated immune responses of a mixed whole-cell and acellular pertussis vaccine schedule in Australian infants: a randomised, double-blind, non-inferiority trial": CONSORT Checklist

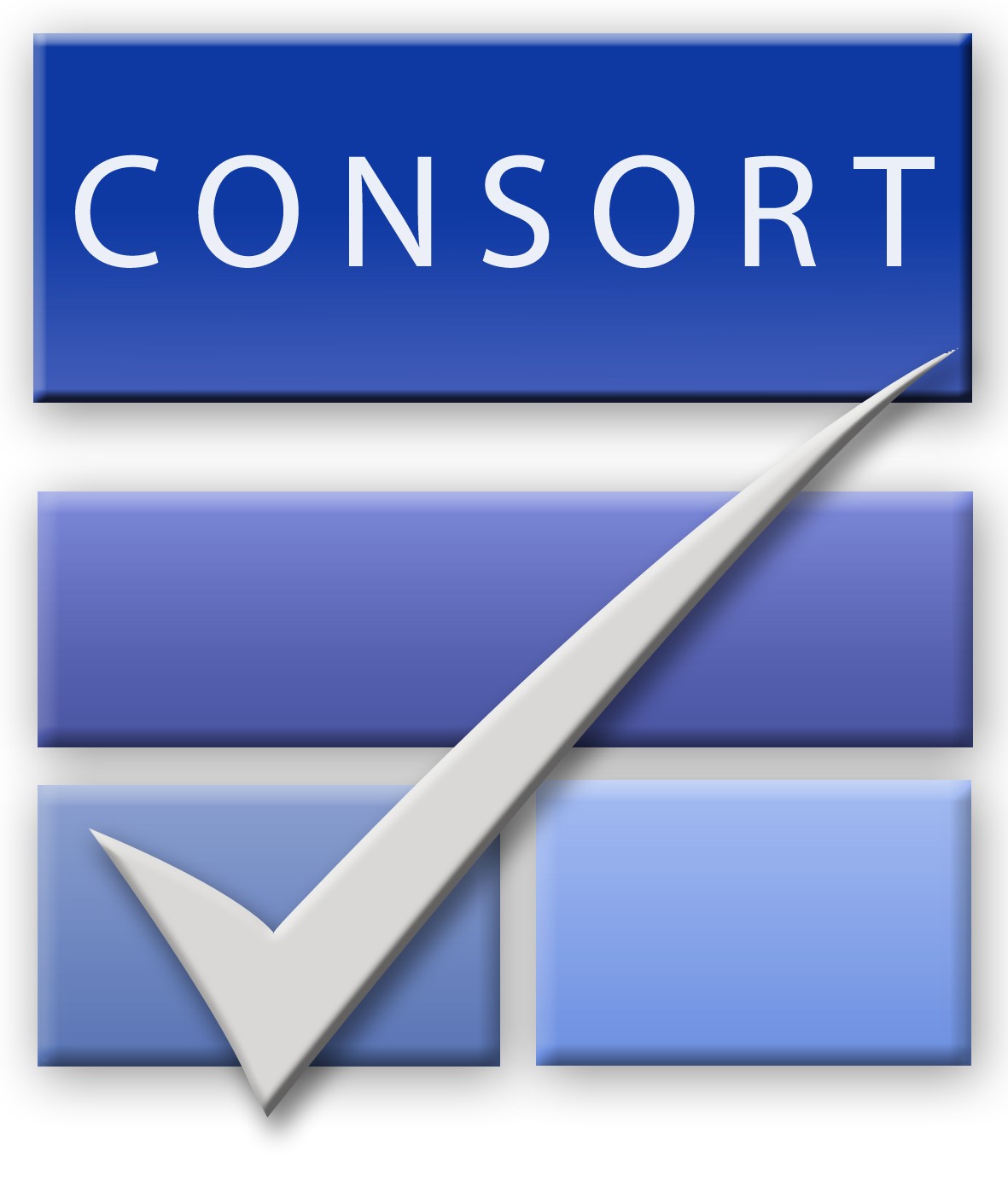
CONSORT 2010 checklist of information to include when reporting a randomised trial*

| Section/Topic | Item No | Checklist item | Reported on page No |
| --- | --- | --- | --- |
| Title and abstract | | | |
|  | 1a | Identification as a randomised trial in the title | Page 1 (identified as non-inferiority trial in the title) |
|  | 1b | Structured summary of trial design, methods, results, and conclusions (for specific guidance see CONSORT for abstracts) | Pages 1 to 4. Hypothesis concerning non-inferiority (eg, non-inferiority of the mixed scheduled compared to the aP-only schedule) and non-inferiority margin are defined in the methods section |
| Introduction | | | |
| Background and objectives | 2a | Scientific background and explanation of rationale | Pages 7 and 8, including the rationale for using a non-inferiority design |
|  | 2b | Specific objectives or hypotheses | Introduction, page 7 |
| Methods | | | |
| Trial design | 3a | Description of trial design (such as parallel, factorial) including allocation ratio | Pages 8 and 9 (study design and participants; randomisation and masking) |
|  | 3b | Important changes to methods after trial commencement (such as eligibility criteria), with reasons | NA |
| Participants | 4a | Eligibility criteria for participants | Page 7 (study design and participants); statistical analysis plan and study protocol |
|  | 4b | Settings and locations where the data were collected | Page 8 (study design and participants) |
| Interventions | 5 | The interventions for each group with sufficient details to allow replication, including how and when they were actually administered | Pages 9 and 10 (randomisation and masking; procedures) |
| Outcomes | 6a | Completely defined pre-specified primary and secondary outcome measures, including how and when they were assessed | Page 9, 10, and 11 (procedures and outcomes) |
|  | 6b | Any changes to trial outcomes after the trial commenced, with reasons | NA |
| Sample size | 7a | How sample size was determined | Statistical analysis plan |
|  | 7b | When applicable, explanation of any interim analyses and stopping guidelines | Stage 1: NA |
| Randomisation: |  |  |  |
| Sequence generation | 8a | Method used to generate the random allocation sequence | Page 9 (randomisation and masking) |
|  | 8b | Type of randomisation; details of any restriction (such as blocking and block size) | Page 9 (randomisation and masking; statistical analysis plan and study protocol) |
| Allocation concealment mechanism | 9 | Mechanism used to implement the random allocation sequence (such as sequentially numbered containers), describing any steps taken to conceal the sequence until interventions were assigned | Page 9 (randomisation and masking; statistical analysis plan and study protocol) |
| Implementation | 10 | Who generated the random allocation sequence, who enrolled participants, and who assigned participants to interventions | Randomisation and masking; participants were enrolled by GPC |
| Blinding | 11a | If done, who was blinded after assignment to interventions (for example, participants, care providers, those assessing outcomes) and how | Page 9 (randomisation and masking) |
|  | 11b | If relevant, description of the similarity of interventions | NA |
| Statistical methods | 12a | Statistical methods used to compare groups for primary and secondary outcomes | Page 12 (statistical analysis) and statistical analysis plan |
|  | 12b | Methods for additional analyses, such as subgroup analyses and adjusted analyses | Page 11 (statistical analysis, adjusted models) |
| Results | | | |
| Participant flow (a diagram is strongly recommended) | 13a | For each group, the numbers of participants who were randomly assigned, received intended treatment, and were analysed for the primary outcome | Figure 1 – Trial profile |
|  | 13b | For each group, losses and exclusions after randomisation, together with reasons | Figure 1 – Trial profile |
| Recruitment | 14a | Dates defining the periods of recruitment and follow-up | Page 14 (results) |
|  | 14b | Why the trial ended or was stopped | Stage 1 was concluded once the last child completed the study. Stage 2 is currently recruiting participants |
| Baseline data | 15 | A table showing baseline demographic and clinical characteristics for each group | Table 1 |
| Numbers analysed | 16 | For each group, number of participants (denominator) included in each analysis and whether the analysis was by original assigned groups | Analyses were by original assigned groups. “Numbers analysed” (ITT and PP analyses sets) are available in the [supplementary document](https://osf.io/4cw6g/?view_only=ccb65276705f4f8daeaeaaeb89d36e91); results (page 20) and Table 3 |
| Outcomes and estimation | 17a | For each primary and secondary outcome, results for each group, and the estimated effect size and its precision (such as 95% confidence interval) | Table 2 and Table 3; [supplementary document](https://osf.io/4cw6g/?view_only=ccb65276705f4f8daeaeaaeb89d36e91) |
|  | 17b | For binary outcomes, presentation of both absolute and relative effect sizes is recommended | All the prespecified analyses are included in the [supplementary document](https://osf.io/4cw6g/?view_only=ccb65276705f4f8daeaeaaeb89d36e91) |
| Ancillary analyses | 18 | Results of any other analyses performed, including subgroup analyses and adjusted analyses, distinguishing pre-specified from exploratory | No exploratory analyses were performed. Unadjusted and adjusted models are detailed in the [supplementary document](https://osf.io/4cw6g/?view_only=ccb65276705f4f8daeaeaaeb89d36e91) |
| Harms | 19 | All important harms or unintended effects in each group (for specific guidance see CONSORT for harms) | NA |
| Discussion | | | |
| Limitations | 20 | Trial limitations, addressing sources of potential bias, imprecision, and, if relevant, multiplicity of analyses | Discussion |
| Generalisability | 21 | Generalisability (external validity, applicability) of the trial findings | Discussion |
| Interpretation | 22 | Interpretation consistent with results, balancing benefits and harms, and considering other relevant evidence | Discussion |
| Other information | | |  |
| Registration | 23 | Registration number and name of trial registry | Abstract and statistical analysis |
| Protocol | 24 | Where the full trial protocol can be accessed, if available | Protocol: 10.1136/bmjopen-2020-042838  Statistical analysis plan: 10.1186/s13063-021-05874-6 |
| Funding | 25 | Sources of funding and other support (such as supply of drugs), role of funders | Role of the funding source |

Citation: Schulz KF, Altman DG, Moher D, for the CONSORT Group. CONSORT 2010 Statement: updated guidelines for reporting parallel group randomised trials. BMC Medicine. 2010;8:18.
© 2010 Schulz et al. This is an Open Access article distributed under the terms of the Creative Commons Attribution License (<http://creativecommons.org/licenses/by/2.0>), which permits unrestricted use, distribution, and reproduction in any medium, provided the original work is properly cited.

*We strongly recommend reading this statement in conjunction with the CONSORT 2010 Explanation and Elaboration for important clarifications on all the items. If relevant, we also recommend reading CONSORT extensions for cluster randomised trials, non-inferiority and equivalence trials, non-pharmacological treatments, herbal interventions, and pragmatic trials. Additional extensions are forthcoming: for those and for up-to-date references relevant to this checklist, see [www.consort-statement.org](http://www.consort-statement.org/).
